# Pandemic impacts on healthcare utilisation: a systematic review

**DOI:** 10.1101/2020.10.26.20219352

**Authors:** Ray Moynihan, Sharon Sanders, Zoe A Michaleff, Anna Scott, Justin Clark, Emma J To, Mark Jones, Eliza Kitchener, Melissa Fox, Minna Johansson, Eddy Lang, Anne Duggan, Ian Scott, Loai Albarqouni

## Abstract

**Objectives:** To determine the extent and nature of changes in utilisation of healthcare services during COVID-19 pandemic.

**Design:** Systematic review

**Eligibility:** Eligible studies compared utilisation of services during COVID-19 pandemic to at least one comparable period in prior years. Services included visits, admissions, diagnostics, and therapeutics. Studies were excluded if from single-centres or studied only COVID-19 patients.

**Data sources:** PubMed, Embase, Cochrane COVID-19 Study Register, and pre-prints were searched, without language restrictions, until August 10, using detailed searches with key concepts including COVID-19, health services and impact.

**Data analysis:** Risk of bias was assessed by adapting ROBINS-I and Cochrane Effective Practice and Organization of Care tool. Results were analysed using descriptive statistics, graphical figures, and narrative synthesis.

**Outcome measures:** Primary outcome was change in service utilisation between pre-pandemic and pandemic periods. Secondary outcome was the change in proportions of users of healthcare services with milder or more severe illness (e.g. triage scores).

**Results:** 3097 unique references were identified, and 81 studies across 20 countries included, reporting on >11 million services pre-pandemic and 6.9 million during pandemic. For the primary outcome, there were 143 estimates of changes, with a median 37% reduction in services overall (interquartile range −51% to −20%), comprising median reductions for visits of 42%(−53% to −32%), admissions, 28%(−40% to −17%), diagnostics, 31%(−53% to −24%), and for therapeutics, 30%(−57% to −19%). Among 35 studies reporting secondary outcomes, there were 60 estimates, with 27(45%) reporting larger reductions in utilisation among people with a milder spectrum of illness, and 33 (55%) reporting no change.

**Conclusions:** Healthcare utilisation decreased by about a third during the pandemic, with considerable variation, and with greater reductions among people with less severe illness. While addressing unmet need remains a priority, studies of health impacts of reductions may help health-systems prioritise higher-value care in the post-pandemic recovery.

**Funding, Study registration:** No funding was required. PROSPERO: CRD42020203729

**Strengths and limitations of this study:**

– The review is the first broad synthesis of global studies of pandemic related changes in utilisation across all categories of healthcare services.
– The review provides novel findings informing design of future studies of pandemic-related changes in utilisation and its impacts.
– Limitations include the possibility of publication bias and the potential of our eligibility criteria to exclude important data sources such as studies in single-centres and unpublished datasets from health systems.
– Heterogenous designs and settings precluding meta-analysis.

## Introduction

As the COVID-19 pandemic continues, many studies have reported major changes in utilisation of healthcare services because of such measures as lockdowns and stay-at-home orders. ^1-3^ These changes include large reductions in services, particularly in places hit hard by the pandemic, but also some selective increases, such as for telemedicine. ^4^ Many people have missed out on much needed care, such as vaccination or life-extending interventions for cancer. ^2,5,6^ A World Health Organization survey found disruption to healthcare services greatest among lower income countries, and there are estimates that reduction of essential maternal and child health interventions may cause more than a million additional child deaths. Concurrently the pandemic may also have resulted in some people being spared unnecessary or inappropriate care with has the potential to cause harm. ^9,10^ The problem of too much medicine is well documented, ^11-17^ and multiple global campaigns are addressing this challenge, such as Choosing Wisely, which is active in more than 20 countries. ^18^ As some nations are forced to do more with less in the post-pandemic period, learning from this “natural experiment” in reduced care may help health systems identify and address unnecessary care, and move towards greater sustainability. ^9,10^

Investigating the impact of changes in healthcare utilisation on health outcomes and costs presents major methodological challenges. First, there are many reasons why people have missed care, including fear of becoming infected while visiting a care facility, inability to access care due to lockdown policies, and suspension and cancellation of services such as elective surgery. Second, disentangling populations who have missed necessary care from those who have avoided unnecessary care requires sensitive and nuanced analysis, with adjustment for multiple potentially confounding variables. For instance, simply showing no adverse outcomes in the short term from missing an episode of care does not prove it was unnecessary. Notwithstanding these challenges, quantifying and characterising the unprecedented recent changes in utilisation, and their impact on health outcomes and costs, may help health systems optimise post-pandemic use of resources.

To this end, we conducted what is, to our knowledge, the first systematic review of studies reporting on pandemic-related changes in overall healthcare utilisation. In undertaking this review, we also sought to inform and optimise the design of future investigations of both the on-going changes in utilisation, and the impacts of this natural experiment with less care on health outcomes and costs.

## Methods

As per a detailed protocol registered on PROSPERO ^19^ and uploaded to the Open Science Framework ^20^ (Supplementary File 1) we found, appraised, and synthesised studies that compared healthcare utilisation during the COVID-19 pandemic with a corresponding pre-pandemic period. Our abstract and full review follow the Preferred Reporting Items for Systematic Reviews and Meta-Analyses (PRISMA) statements. ^21,22^ (Supplementary File 2)

### Eligibility Criteria and Search Strategy

#### Inclusion and exclusion criteria

We included studies which compared utilisation of healthcare services over a period of time during the pandemic, as defined by their authors, (the intervention) with a corresponding period at least one year before the pandemic, (the comparator). Healthcare service utilisation included but was not limited to visits or presentations, admissions or hospitalisations, diagnostic services, and therapeutic or preventive interventions. Letters or pre-prints were included if providing enough data for extraction. We excluded surveys of practitioners, studies reporting only on utilisation by patients diagnosed with COVID-19, studies reporting utilisation data for less than one week, from a single centre only, or for non-medical allied health services, and modelling studies that predicted impacts on utilisation.

#### Outcome measures

The primary outcome was the change in utilisation of a healthcare service – such as a visit to a hospital or receipt of diagnostic imaging – between the pre-pandemic and pandemic periods, expressed as a change in absolute numbers and/or percentage change. The secondary outcome was change in the proportions of people using the service, across different levels of disease severity, as reported by authors of the primary study, using for example a triage score.

#### Data sources, searches, screening

We searched PubMed, Embase, the Cochrane COVID-19 Study Register, and pre-print servers via Europe PMC, from inception until 10^th^ August, 2020, with search strings that included the following broad concepts: COVID-19, health services, admissions, and impact. (Supplementary File 3) No restrictions by language were imposed. Following screening of articles for inclusion, we conducted a backwards (cited) and forwards (citing) citation analysis in Scopus/Web of Science on all included studies, and additional articles were screened for inclusion. We also consulted experts for other public reports.

Pairs of review authors (RM, SS, ZM, AS, JC, EK, ET, LA) independently screened the titles and abstracts against the inclusion criteria, and repeated the process following full-text retrieval. Any screening disagreements were resolved by discussion, or reference to a third author (RM or LA). A list of studies in single centres, excluded at screening stage, was recorded and is available on request from authors.

### Data Collection and Analysis

#### Data extraction

Pairs of authors (RM, SS, ZM, AS, ET, LA) independently extracted data from included studies and resolved discrepancies, with referral, as necessary, to a third author (LA, RM). We developed, piloted, and used a data extraction form in Microsoft Excel for study characteristics and outcome data. We extracted data on study location, design, setting, (e.g. hospital) pandemic period and comparator, and primary and secondary outcomes.

Pairs of review authors (RM, SS, ZM, AS, LA, ET) independently assessed the risk of bias for each included study using a risk of bias tool adapted from the ROBINS-I tool ^23,24^ as per guidance provided by Cochrane for assessing risk of bias in uncontrolled before-after studies including interrupted time series, ^23^ and a tool developed by the Cochrane Effective Practice and Organization of Care group. ^25^ All disagreements were resolved by discussion or referral to a third author (RM, LA, SS). The domains assessed included bias related to: confounding (a. the possibility that extraneous events occurring around the time of the pandemic may have influenced the outcome, b. how well the study accounted for pre-intervention trends in utilisation); selection of participants; outcome measurement; and selective reporting of results. (Supplementary file 4) Each potential source of bias was graded as low, high or unclear, with the exception of grading for the pre-intervention trends, which was graded as low, moderate or high.

#### Data synthesis and analysis

As anticipated in the protocol, the considerable clinical and statistical heterogeneity in settings, outcome measures, and methods precluded a formal quantitative meta-analysis. Hence, we summarised the results using descriptive statistics (percentage change expressed as median and interquartile range), graphical figures and a narrative synthesis. In line with the “Synthesis without meta-analysis (SWiM) in systematic reviews: reporting guideline” ^26^ we summarised findings for the primary outcome grouped by four service types: visits or presentations; admissions or hospitalisations; diagnostic or imaging investigations; and therapeutic or preventive interventions.

For the secondary outcome, we developed and report three categories which relied on the indicators of disease severity employed by primary study authors: a larger or smaller reduction among those with milder forms of illness, compared to people with more severe forms of illness; and no change. An example of a secondary outcome for a study of emergency department, ED, visits would be the triage scores, used to assess severity of those attending. Two authors (RM, LA) independently assigned a category for each secondary outcome, informed where possible by statistics provided in primary studies, with oversight and resolution of any discrepancies from within the clinical authorship team, (IS, EL, MJ).

As per details in the protocol, we planned to conduct a limited meta-analysis and sensitivity analysis in situations where there was a sufficient number of clinically and statistically homogeneous studies. Also, as per protocol, we restricted our analysis to data in the primary studies, rather than correlating findings with external information, such as stages of lockdown.

### Patient and public involvement

The chief executive officer of a peak state-based consumer health organisation had input into the interpretation of the review data, and the revising and approval of the draft manuscript.

### Ethics

No ethics approval was required.

### Changes from protocol

Several minor changes comprised: during data extraction we could not confidently assess whether each utilised service was not provided or just deferred; finalisation of the adapted tool for risk of bias resulted in five domains, not six (two domains related to outcome measurement were combined), with one domain assessed as low, moderate, high, rather than unclear, low and high, with each grade supported by a comment; and given the very large number of included studies, we included data from studies reporting only a percentage change in service utilisation, without contacting authors requesting the absolute numbers.

## Results

### Study selection

We identified 4817 records through electronic database searching, 323 more through forward-backward citation analysis, and one from other sources, for a total of 3097 unique records. After screening titles and abstracts, we excluded 2929 records, and selected 179 records for full-text screening, of which 98 were excluded with reasons recorded. This left 81 studies which were included in the review. (Figure 1)

**Figure 1.**
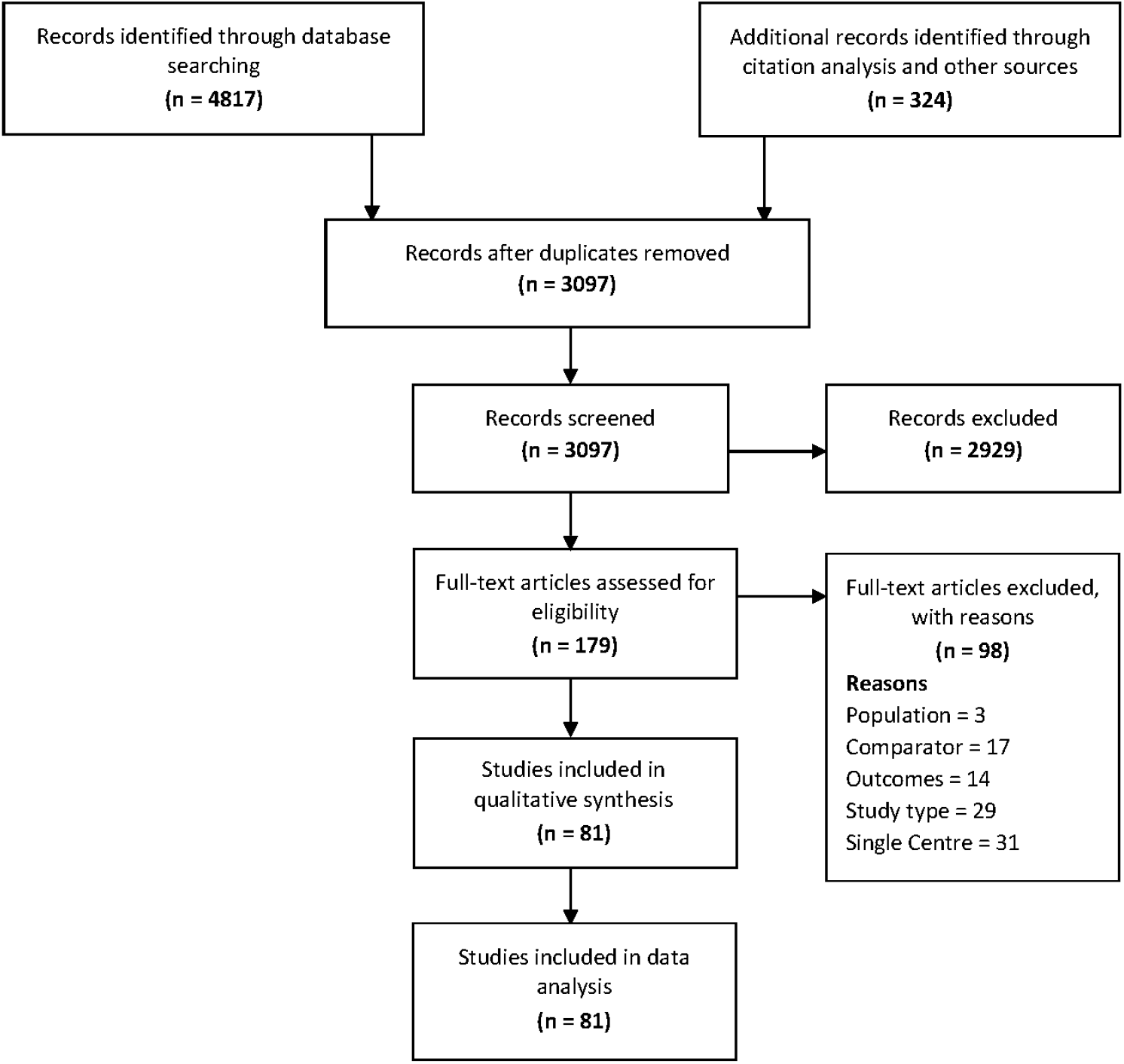
PRISMA Flow Diagram.

### Characteristics of included studies

The 81 included studies collectively report on more than 6.9 million in the pandemic and over 11 million in the comparator pre-pandemic period. Studies reported across multiple locations: 3 were multi-national; 20 originated from the United States (US); 15 from Italy; 8 from France; 6 from Germany; 5 from the United Kingdom; 3 from Spain; 2 from each of Taiwan, Hong Kong, Greece, Denmark, Qatar, Australia; and 1 from each of Argentina, China, Canada, Brazil, Belgium, Chile, Monaco, Turkey, and Portugal. Four studies were from low-or middle-income countries. The healthcare setting were: hospitals only (41; 51%); both ED and hospitals (12; 15%); ED only (15;19%); and primary care and/or community (9;11%). More than one third of studies reported on healthcare services related to cardiovascular diseases (n=33; 41%); 14 (17%) to emergency services; 12 (15%) to general services such as immunization and primary care; and 22 (27%) on services related to different conditions including orthopaedic and trauma services, gastroenterology, and mental health. Of the included studies, 14 (17%) were national studies and 9 (11%) used time-trend data (Table 1; Supplementary file 5).

**Table 1.** Summary characteristics of included studies (n=81)

| Characteristics of included studies | No (%) |
| --- | --- |
| **Scope** |  |
| National | 14 (17%) |
| Multi-centre | 67 (83%) |
| **Disease categories** |  |
| Cardiovascular | 33 (41%) |
| Emergency Services (adult and paediatric) | 14 (17%) |
| General (including vaccination and hospice) | 12 (15%) |
| Digestive | 5 (6%) |
| Orthopaedic and Trauma | 5 (6%) |
| Others (e.g. mental health, urology, neurology) | 12 (15%) |
| **Setting** |  |
| Hospitals (or inpatient care) | 41 (51%) |
| Emergency | 15 (19%) |
| Emergency and Hospital | 12 (15%) |
| Community and/or outpatient | 9 (11%) |
| Hospital and outpatient | 4 (5%) |
| **Study design\*** |  |
| **Time trend** |  |
| Time trend – multiple prior year | 5 (6%) |
| Time trend – single prior year | 4 (5%) |
| **Same period (before – after)** |  |
| Same period – multiple prior year | 16 (20%) |
| Same period – single prior year | 56 (69%) |
| **Country** |  |
| Multi-national | 3 (4%) |
| Americas | 24 (30%) |
| Europe | 45 (56%) |
| Asia & Australia | 9 (11%) |

\*This refer to the type of data
used in included studies rather than the type of analysis

### Risk of bias assessment

For the majority of studies there was insufficient information on which to judge the possibility that extraneous events occurring between pre-pandemic and pandemic periods may have influenced healthcare utilisation, or to assess the risk of bias arising from differences between those eligible to utilise healthcare services in the pre-pandemic and pandemic periods (76/81; 94%). 69% (56/81) of studies were considered to be at high risk of bias due to insufficient data for characterising pre-pandemic utilisation. In contrast, three studies (4%) were judged to be at low risk of bias on this domain due to adequate data and analysis to permit characterisation of pre-pandemic trends in utilisation. 63% (51/81) of studies were judged to be at high or unclear risk of bias from using different methods used to assess utilisation in the pre-pandemic and pandemic period, or lacking information on which to judge this domain. Most studies (n= 74; 91%) were judged to be at low risk of bias in selective reporting of results.

### Main findings

The 81 studies reported 143 estimates of changes in healthcare utilisation between pandemic and pre-pandemic periods, of which 136 (95.1%) were a reduction. The percentage change ranged between a 49% increase and an 87% decrease with a median 37.2% reduction (interquartile range −50.5% to −19.8%). For the 64 estimates about changes in cardiovascular service utilisation, from 33 studies, the median reduction was 29.3% (−41.3% to −17%). For the 13 estimates from the 9 studies using time-trend data, the median reduction was 37.3% (−45% to −25.2%). For all studies, the weekly median percentage changes starting from mid-February until late May 2020 are plotted graphically in Figure 3, showing greatest reductions through March and April. (Full data in Supplementary file 5)

**Figure 2.**
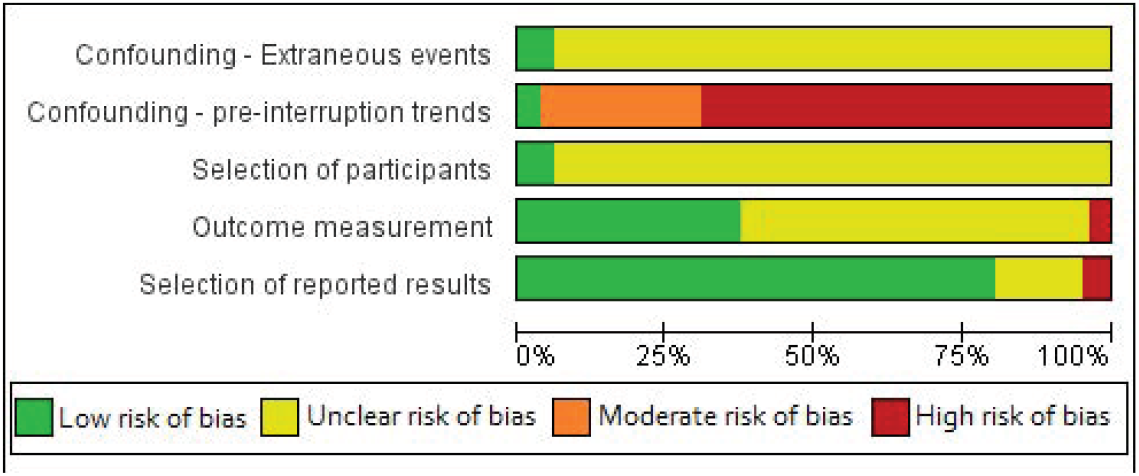
Summary of Risk of Bias Assessments.

**Figure 3.**
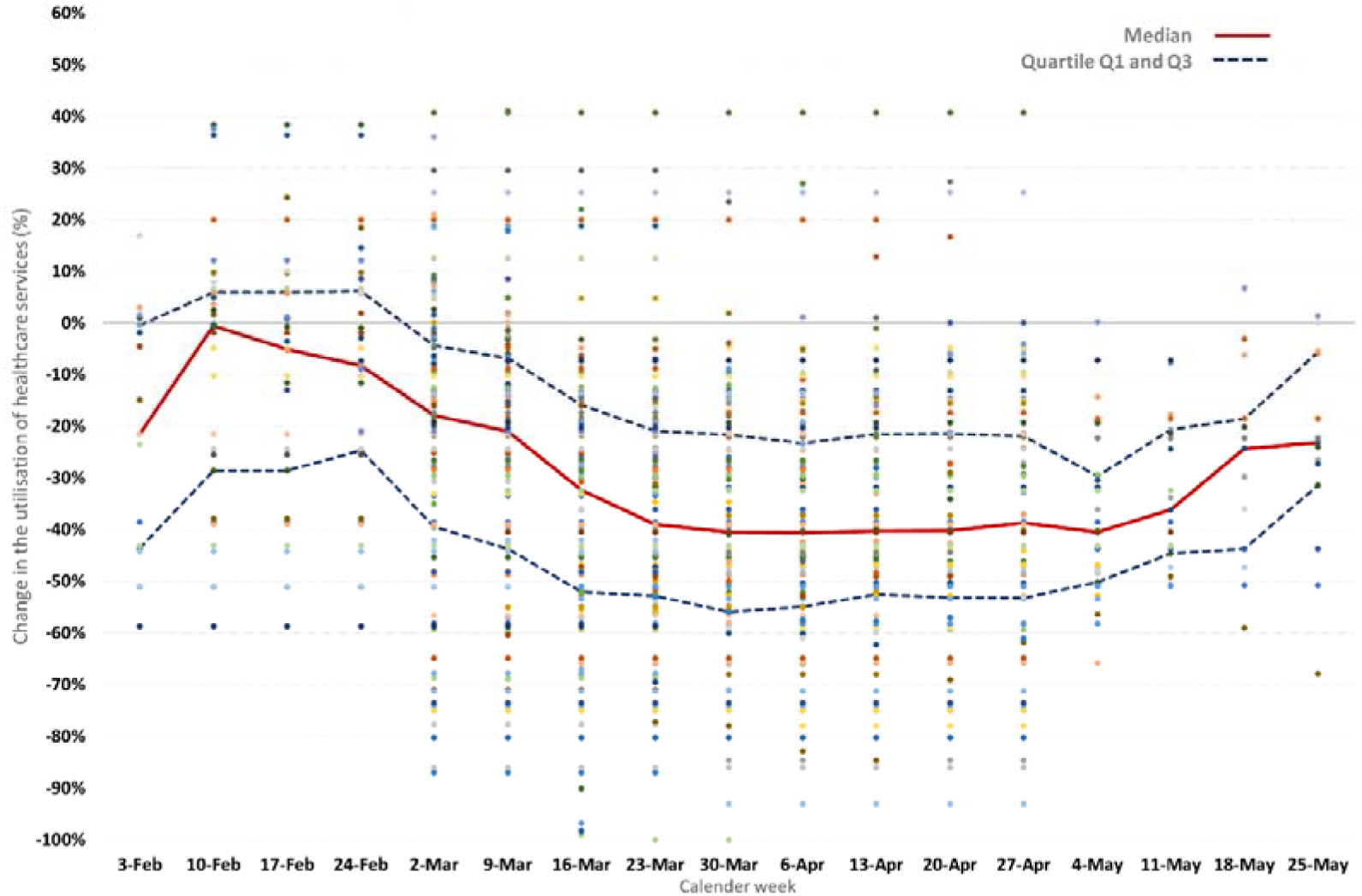
Pandemic related changes in healthcare utilisation.

We categorized the 143 estimates of change into 4 groups according to the type of healthcare service: 41 estimates for healthcare visits; 43 estimates for admissions; 12 estimates for diagnostics (e.g. imaging, pathology, screening investigations); and 47 estimates for therapeutics (e.g. surgery, vaccinations). All medians are reported in Table 2, with results of individual studies reported in Supplementary file 5.

**Table 2.**
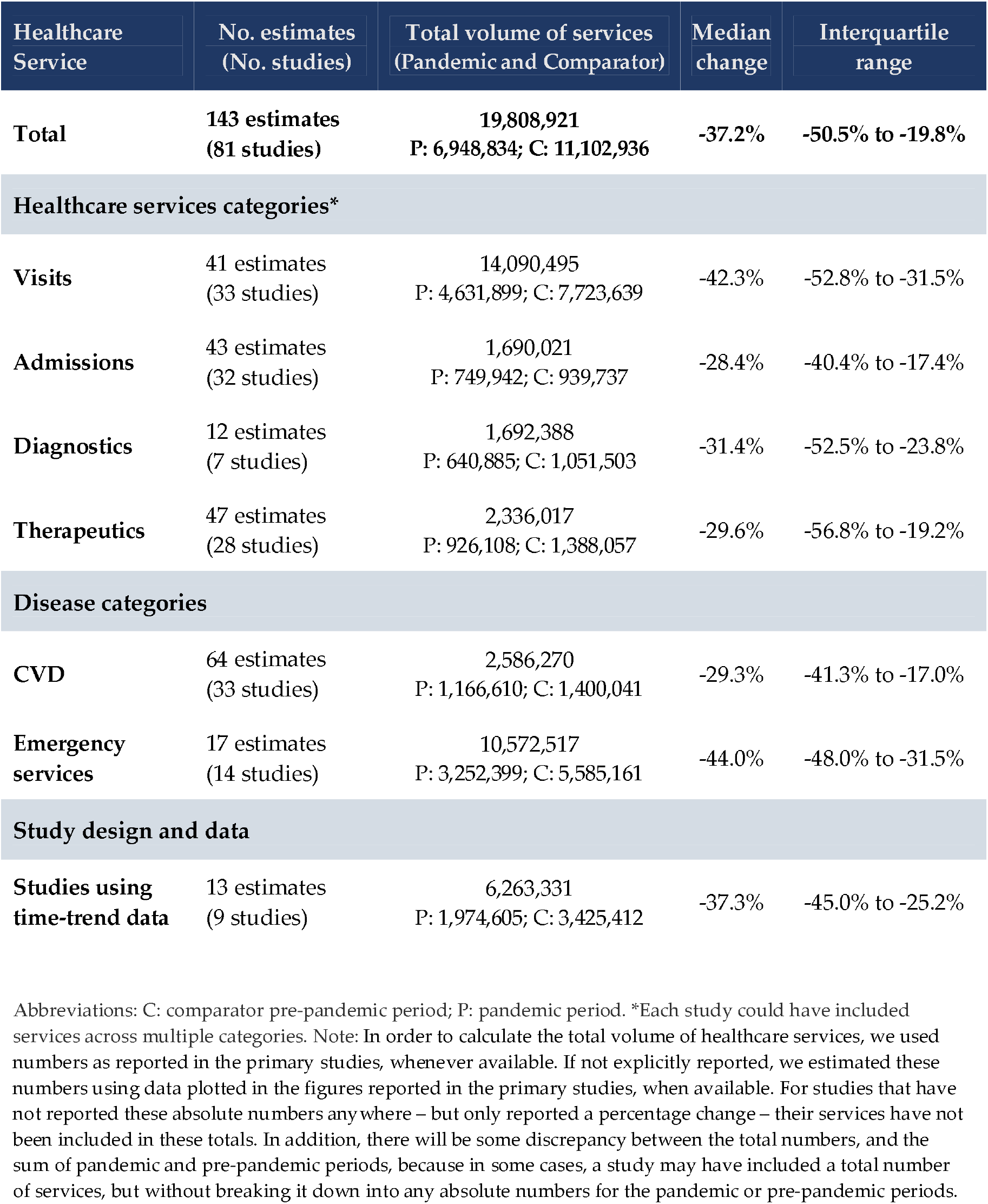
Median changes in utilisation across categories of healthcare services.

### Changes in visits

The percentage change for healthcare visits or presentations ranged between a 49% increase and an 86% decrease, with a median 42.3% reduction (−52.8% to −31.5%). Major reductions in visits to EDs were seen in multiple studies, such as a large national US study from the Centres for Disease Control and Prevention reporting a 42% reduction during April, rising to a 26% reduction at the end of May, compared to 2019. ^1^ That study found the largest absolute reduction involved people presenting with abdominal pain, with over 66,000 fewer ED visits per week for this complaint during April. In terms of age group, the largest reduction (−72%) was seen for children 10 years and under. ^1^ A metanalysis of a subgroup of six studies of ED visits that reported adequate data for meta-analysis (effect estimates and 95% CIs) was attempted, but demonstrated considerable heterogeneity (I^2^ >95%).

### Changes in admissions

The percentage change in the number of admissions ranged between a 20% increase and an 87% decrease, with a median 28.4% reduction (−40.4% to −17.4%). For example, a large study of the weekly admission rates for acute coronary syndrome in England showed a substantial reduction by the end of March (−40%) which partly rebounded by the last week of May 2020, (−16%). ^27^

### Changes in diagnostics

The percentage reduction ranged between 10% and 85%, with a median 31.4% reduction (−52.5% to −23.8%); no study reported any increase in the use of diagnostic and imaging procedures. The magnitude of reductions in diagnostic tests and imaging followed a trend over time similar to those observed in the previous categories, but with a far smaller number of estimates. (See Figures 5.4a-d, Supplementary file 5) For example, a study of imaging case volumes within the largest healthcare system in New York State found a 28% reduction in imaging volumes for March to mid-April 2020 across all locations and imaging modalities, ^28^ while a separate US study found volumes recovering through late April, but still 36% lower in the third week of May, compared to 2019. ^29^

### Changes in therapeutics

The percentage change in therapeutic and preventive care ranged between a 27% increase and an 80% decrease, with a median 29.6% reduction (−56.8% to −19.2%). For example a large study of routine childhood vaccination in England found fewer children receiving the first MMR dose, with a reduction of 24% in the final week of March, which rose to a 27% increase in the third week of April, compared to the same period in 2019. ^5^

### Secondary Outcome

Thirty-eight of the included studies reported a total of 60 secondary outcomes relating to potential changes in healthcare utilisation according to the disease severity of the service user. Despite the considerable heterogeneity in settings and services, for almost half of these outcomes, (27 of 60; 45%) we observed a pattern of larger reductions in utilisation among those with milder or less severe illness compared to those with more severe disease. For 33 of 60 outcomes (55%) there was no change. (Figure 4)

**Figure 4.**
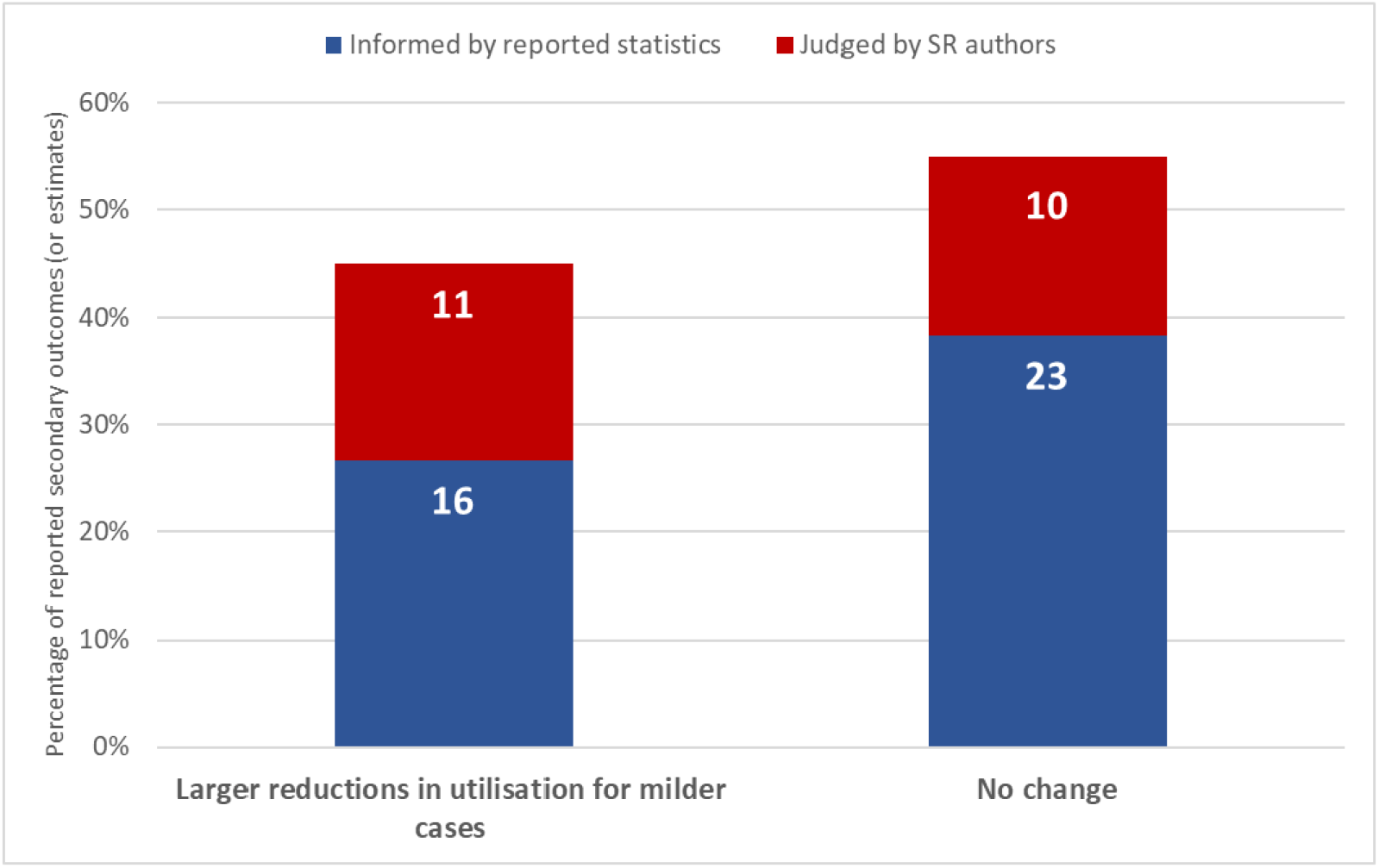
Differential reductions in utilisation relating to severity.

A national Italian study of urgent endoscopy reported a 40% reduction in utilisation overall, with bigger reductions in the proportion of patients with a negative finding on upper endoscopy between pre-pandemic and pandemic periods.^3^ A study of three psychiatric emergency services in Paris found a 55% overall reduction in presentations in the first 4 weeks of lockdown, with greater reductions for consultations for anxiety and stress, and smaller reductions for consultations for psychotic disorders. ^30^ Authors speculated that “some people may find new strengths and coping strategies during disasters” and “the current results may arise from an elevation in resilience.” Most strikingly, multiple studies reporting reduced acute coronary syndrome presentations found these reductions were much greater for the less severe non-ST-segment elevation myocardial infarction (NSTEMI) events compared to ST-segment elevation myocardial infarctions, (STEMIs). ^27, 31^ An example is a large English study reporting reductions in admissions of 42% for NSTEMI events versus 23% for STEMI. ^27^ In contrast, other studies found no change in presentations according to severity, including a national Portuguese study reporting a 48% reduction in ED episodes – from an expected 570 000 to an observed 295 000 in March 2020 – but no significant change in proportions of different triage categories. ^32^

## Discussion

This review of 81 studies involving over 19.8 million services provided across 20 countries found consistent evidence of major reductions in the utilisation of healthcare services during the pandemic period up to May 2020, compared to previous years, despite some studies reporting increases. Although a meta-analysis was not possible, we found a median reduction of 37% of services overall, which was highest for visits (42%) and slightly lower for admissions (28%), diagnostics (31%) and therapeutics (30%). Many studies also found larger reductions in utilisation among populations with milder or less severe illness. Few studies were assessed as having a low risk of bias, with lowest risk of bias for studies using time-trend data to establish trends in the years leading up to 2020. For the 9 studies using time-trends, the median reduction in utilisation was 37%.

Our review has several strengths. First, we synthesized the most recent data reported in primary studies up to the end of May 2020, which corresponds to the peak of the pandemic in many countries, and provides a baseline for longer-term data on on-going changes in utilisation and the cumulative deficit of care. Second, the review constitutes the first broad synthesis of global studies of pandemic related changes in utilisation across all categories of healthcare services. Third, the review adhered to rigorous Cochrane, ^24^ PRISMA ^21, 22^ and SWiM ^26^ standards. Study limitations include the inability to undertake a meta-analysis because of considerable heterogeneity, the possibility of publication bias, the potential of our eligibility criteria to exclude important data sources such as studies in single-centres and unpublished datasets from health systems, subjectivity in our assessments of the secondary outcomes, and the use of an adapted but unvalidated risk of bias tool.

The massive global reduction in healthcare utilisation summarised in this review makes a compelling case for prioritising efforts that address the unmet needs of those with non-COVID 19 illness. Consistent messages from the primary studies include calls for monitoring the long-term impacts of this missed care, public campaigns to urge people to seek medical care when they need it, and better preparedness for reducing the extent of missed care in future waves of the pandemic. Evidence of excess population mortality, in addition to deaths from COVID-19, and related phenomena such as increases in out-of-hospital cardiac arrests and contacts with emergency phone-lines ^33,34^ make these calls to action even more urgent. Conversely, the review’s finding that reductions often tended to be greater for milder or less severe forms of illness, combined with existing evidence about too much medicine, ^11-17^ suggest that for some people, missing care may not have caused harm.

This unprecedented pandemic-induced natural experiment in reduced healthcare utilisation provides a genuine opportunity to learn more about what services populations and healthcare systems came to regard as lesser priorities, when redistribution of resources towards more essential services was needed to minimize mortality in a crisis. As others have suggested, ^35,36^ greatly reduced ED attendances around the world for non-urgent complaints indicate an opportunity to inform and implement new strategies and models of care that maximise the appropriateness of visits in the future. Even at the heart and height of the epidemic in Northern Italy, in paediatric EDs doctors found reductions in the mildest presentations accounted for more of the decrease in overall presentations, suggesting that “most of the non-relevant pathologies usually seen at our EDs have been avoided” thus freeing resources to “provide critical services to patients suffering from medical emergencies in a timely manner.” ^36^ Our review adds weight to the view that the post-pandemic recovery provides a rare window of opportunity for systematic changes in healthcare systems aimed at reducing low-value care, including overtreatment and overdiagnosis. ^9,10,37^

Many questions about the causes and impacts of the changes in healthcare utilisation documented in our review call for careful analysis and further research. (See Box 1) High quality time trend analyses are needed to better understand the extent and nature of on-going changes in utilisation, as are long-term cohort studies for collecting patient-centred outcomes to assess impacts on health, costs, and equity. Consultations with consumers during the pandemic highlight the need to understand how the pandemic may differentially impact the most vulnerable, and the need to prioritise those at most need. ^38,39^ Rigorous qualitative research investigating people’s experience of avoiding or missing care, and professional responses to changes in process and practice, will also be important. We found no study which explicitly examined changes in utilisation of low-value healthcare services, which warrants further research. The extent and effects of substitution, such as with telehealth or self-care also requires investigation. Experience with SARS almost 20 years ago revealed significant drops in healthcare service utilisation in the most affected regions ^40^ and long periods before some rates returned to baseline. ^41^ Given the growing evidence about unnecessary care since then, it may be more beneficial for populations and their health systems if utilisation rates of some services do not return to pre-pandemic levels. Addressing genuine unmet need and winding back the harm and waste of unnecessary care are not conflicting interests, but rather two sides of a coherent strategy to efficiently improve human health.

**Box 1:**
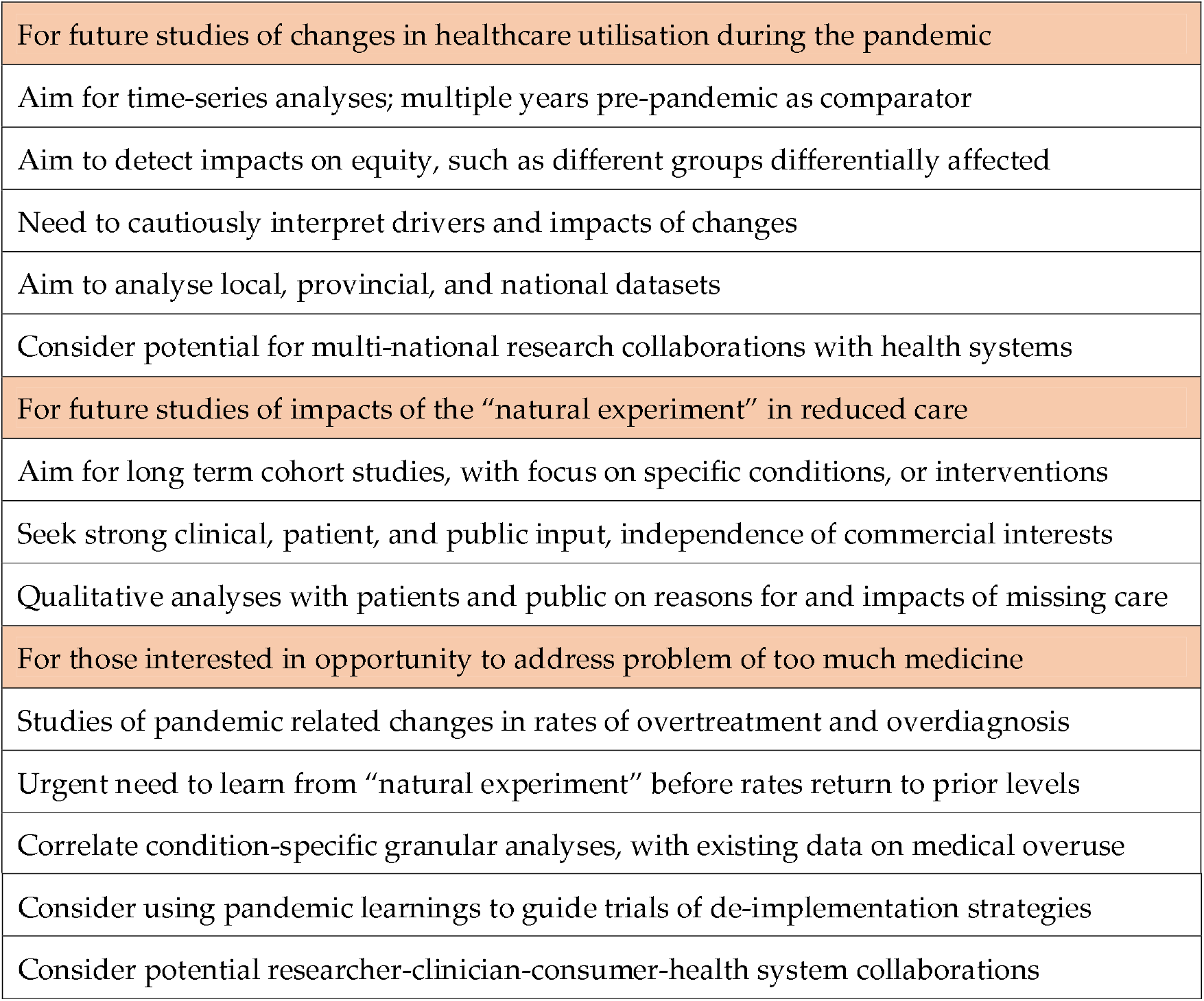
Future Research.

#### Summary Box

##### Section 1: What is already known on this topic

- Multiple primary studies have reported reductions in utilisation of various healthcare services during the COVID-19 pandemic

##### Section 2: What this study adds

- This systematic review is the first to quantify and characterise reductions in health service utilisation on a global scale, across countries, settings, and service types
- The review identifies major reductions in use of services across 20 nations, with a median reduction of 37% overall and reductions of similar magnitude across key service categories of visits, admissions, diagnostics, and therapeutics
- Importantly, reductions in utilisation have tended to be greater among populations with milder or less severe symptoms or conditions
- While controlling the COVID-19 pandemic and tackling unmet needs of those with non-COVID illness remain priorities, examining changes in utilisation may also offer learnings on identifying and reducing unnecessary care in the post-pandemic recovery

## Data Availability

We have provided all data about all included studies, and a list of those studies, in the Supplementary files.

## Acknowledgements

Thanks to Paul Glasziou, Kim Sutherland and Karsten Jorgensen for comments on a draft of this manuscript.

## Contributor and Guarantor

Conception/design: RM, LA, SS, ZM,AS, JC,MJ; Acquisition, analysis or interpretation of data: RM, SS, ZM, AS, JC, ET, MJ, EK, MF, MJ, EL, AD, IS, LA; First draft of manuscript: RM, LA; Manuscript drafting, revision, approval: RM, SS, ZM, AS, JC, ET, MJ, EK, MF, MJ, EL, AD, IS, LA. Overall guarantors: RM, LA. The guarantor accepts full responsibility for the work and/or the conduct of the study, had access to the data, and controlled the decision to publish. The corresponding author attests that all listed authors meet authorship criteria and that no others meeting the criteria have been omitted.

## Copyright/Licence for Publication

The Corresponding Author has the right to grant on behalf of all authors and does grant on behalf of all authors, a worldwide licence to the Publishers and its licensees in perpetuity, in all forms, formats and media (whether known now or created in the future), to i) publish, reproduce, distribute, display and store the Contribution, ii) translate the Contribution into other languages, create adaptations, reprints, include within collections and create summaries, extracts and/or, abstracts of the Contribution, iii) create any other derivative work(s) based on the Contribution, iv) to exploit all subsidiary rights in the Contribution, v) the inclusion of electronic links from the Contribution to third party material where-ever it may be located; and, vi) licence any third party to do any or all of the above.

## Patient and Public Involvement

The most senior officer from a state peak consumer health organisation is a co-author on this review and was involved in the study before the protocol was finalised. The consumer representative provided feedback on the protocol and draft manuscripts, was consulted during the process of the review, was involved with interpretation of results, and will advise on methods for dissemination of study results to the public.

## Transparency Statement

The lead and senior authors RM and LA affirm that the manuscript is an honest, accurate, and transparent account of the study being reported; that no important aspects of the study have been omitted; and that any discrepancies from the study protocol as originally planned and registered have been explained in a special section in the Methods.

