## Supplementary file for "Pandemic impacts on healthcare utilisation: a systematic review"

### Supplementary file 1

#### Pandemic changes in healthcare utilisation: a protocol for a systematic review

Moynihan R [1], Sanders S [1], Michaleff Z [1], Scott AM [1], Clark J[1], Fox M[2], Duggan A[3], Lang E[4], Johansson M[5], Scott I[6], Kitchener E[7], To E[8], Albarqouni L.[1]

1. Institute for Evidence-Based Healthcare, Faculty of Health Sciences and Medicine Bond University, Gold Coast, Australia;
2. Health Consumers Queensland, Brisbane, Queensland, Australia;
3. Australian Commission on Safety and Quality in Healthcare, Sydney, Australia;
4. Cumming School of Medicine, University of Calgary, Alberta Health Services, Calgary, Canada;
5. Cochrane Sustainable Healthcare, Sweden;
6. Princess Alexander Hospital & The University of Queensland;
7. MPH Student, Griffith University, Queensland, Australia;
8. Medical student, University of Calgary, Alberta, Canada;

Physical address for corresponding author, Ray Moynihan: Institute for Evidence-Based Healthcare, Faculty of Health Sciences and Medicine Bond University, 14 University Drive, Robina, Gold Coast, Australia, 4226.

##### BACKGROUND

As the covid-19 pandemic continues, increasing numbers of studies are reporting major changes in utilisation of healthcare services, including large drops in services during certain periods,[1-3] as well as some increases, such as the use of telemedicine.[4] While many people have missed much needed care, such as vaccination or life-saving interventions,[2] others may be avoiding unnecessary or inappropriate care which would have caused them more harm than good.[3] A large and growing evidence base suggests the problem of too much medicine is widespread, including low value care which may carry no benefit, and overdiagnosis, which can cause more harm than good. [5-11] Multiple global campaigns are attempting to address this challenge, such as Choosing Wisely, which is active in more than 20 nations.[12] As nations are forced to do more with less, post-pandemic, learning from this "natural experiment" in less care may help health systems address the challenges of unnecessary care, and move towards more sustainability.[13,14]

Understanding the impact of these large changes in healthcare utilisation, on health outcomes and costs, will present a great methodological challenge. First, there are many

[1] non-first/last authors are indicative order only

reasons why people have missed care, including fear of visiting hospitals during the pandemic, inability to visit due to lockdown circumstances, or the unavailability of a service such as suspended elective surgery. Second, disentangling those groups who have missed needed care, from those who have avoided unnecessary care, will require sensitive and sophisticated analysis, considering multiple potentially confounding variables. Moreover, simply showing no adverse outcomes from missed care – such as a missed visit to a general practitioner – does not automatically mean that episode of missed care was unnecessary. Notwithstanding these challenges, understanding the unprecedented recent changes in utilisation and their impact, may help health systems, and the societies which fund them, optimise resource-use post-pandemic.

As a first step to that understanding, we aim to conduct a systematic review of studies which have reported on pandemic-induced changes in healthcare utilisation. We aim to examine the extent and nature of changes, particularly any reported changes in the severity of symptoms of people seeking or receiving care.[3] The broader purpose is to inform any future investigations of the impact of this natural experiment in less care on health outcomes and costs.

#### METHODS

We aim to find, appraise, and synthesise studies that assessed the impact of the covid-19 pandemic on the utilisation of healthcare services, compared to a corresponding period of time prior to the pandemic. This systematic review will be reported following the Preferred Reporting Items for Systematic Reviews and Meta-Analyses (PRISMA) statement.[15] The review protocol was developed prospectively and was registered on the Open Science Framework (https://osf.io/) and on Prospero ( https://www.crd.york.ac.uk/prospero/). We will also follow the "2 week systematic review" (2weekSR) processes for this review.[16] In relation to the PICO for this systematic review, the P will be a population of people seeking or using a service within the healthcare system, the I will be the pandemic period as defined by primary study authors, the C will be a comparable period at least one year prior to the study period, and the O will be change in utilisation (primary outcome) and change in disease severity of the people using the service, (secondary outcome).

##### Studies to be included

###### *Population*

We will include studies that report changes in the utilisation of healthcare services by patients and public, irrespective of age. We will exclude studies that reported on the utilisation of healthcare services by patients diagnosed with covid-19.

###### *Interventions and Comparators*

We will include studies which compare utilisation during any period within the pandemic, with a similar period in at least one year before the pandemic. We will therefore include studies which compare – for example – April 2019 utilisation with April 2020 utilisation, but due to concerns about reliable comparisons, we will exclude studies which use the immediate pre-pandemic period as a comparator, (e.g. November 2019). We will include studies which report data from national or regional sources, of more than one centre, so we will exclude studies within a single unit or single hospital, due to limitations on generalisability.

*Outcomes*
The primary outcome is the extent of changes in utilisation of a healthcare service between the pre-pandemic comparison period and the pandemic period. Healthcare service will include but not be limited to *consultation healthcare services* such as presentations or admissions to hospitals or visits to primary care; *diagnostic healthcare services* such as diagnostic imaging/investigations, laboratory testing; and *therapeutic or preventive healthcare services* such as prescriptions, or surgeries or utilisation of vaccinations. These healthcare services can be broad and may include packages of, rather than single isolated, healthcare services. Therefore, in the case of a broad package, the primary outcome for the purposes of our review will be the initial indication for the healthcare services utilisation, if that data is available in the primary study, (e.g. admission due to a stroke is an initial indication for a subsequent series of healthcare services including diagnostic investigations and therapeutic services).

The secondary outcome is the nature of the changes in relation to the people using the service, specifically changes in disease severity or diagnostic spectrum, (e.g. any changes in proportions of patients with mild or severe illness).

We will exclude studies which report utilisation for a time period less than one week in duration, because of the brevity of the time period, and the possibility of differences on different days of the week. We will exclude studies which do not include data on changes in routine healthcare utilisation, but rather only describe changes in healthcare processes, incidence/prevalence of conditions/diseases only, the nature of new practices, or the impacts of covid-19 on individual patients. We will exclude non-medical allied health services.

*Study design*
We will include any observational studies using clinical, hospital or health system administrative data and/or medical records reporting utilisation in a period after the pandemic was declared, and at least one corresponding period in the years prior to the pandemic. This will include before-after studies and interrupted time series studies. We will exclude surveys of healthcare practitioners, cross-sectional studies, any trials, or studies using modelling to predict impacts on utilisation.

*Rational for selection and prioritisation of outcomes*
We selected and prioritised the outcomes based on (i) a review of the outcomes reported in a sample of potentially included studies collected before the Systematic Review by 2 review authors (RM, LA); (ii) a discussion among the whole review team, which includes clinical advisors, methodological experts, and a patient and public (consumer) representative. Primary and secondary outcomes directly address the Systematic Review question, which is investigating the extent and nature of changes in healthcare utilisation due to the pandemic.

#### Search strategies to identify studies

##### Database search strings

We will search PubMed, Embase and the Cochrane COVID-19 Study Register and pre-print servers via Europe PMC, from inception until Monday 10th August, 2020, with an update close to date of submission. We designed a search string in pubmed that included the following concepts: Covid-19 AND Health services AND Admissions AND Impact. This search string was translated for use in other databases using the Polyglot Search Translator.[17] The complete search strings for all databases are provided in Appendix 1.

##### Restriction on publication type

No restrictions by language or publication date will be imposed. We will include publications that were published in full, as well as letters, or pre-prints, where data on the primary outcome is sufficient for data extraction. We will seek expert advice on the existence of other public reports unavailable in peer-reviewed journals and they will be included if all inclusion criteria are met.

##### Other searches

We will conduct a backwards (cited) and forwards (citing) citation analysis in Scopus/Web of Science on the included studies identified by the database searches, and these will be screened against the inclusion criteria.

#### Study selection and screening

Pairs of review authors [RM, SS, ZM, AS, JC, EK, ET, LA] will independently screen the titles and abstracts in Endnote for inclusion against the inclusion criteria. One review author [JC] will retrieve full-text, and pairs of authors [RM, SS, ZM, AS, JC, EK, ET, LA] will screen the full-texts for inclusion. Any screening disagreements will be resolved by discussion, or reference to a third author [RM or LA]. The selection process will be recorded in sufficient detail to complete a PRISMA flow diagram and a list of excluded (full-text) studies with reasons for exclusions. A list of studies in single-centres, excluded at title and abstract screening stage, but which otherwise meet inclusion criteria, will be recorded and made available on request from authors.

#### Data extraction

We will develop and use a data extraction form for study characteristics and outcome data, which will be piloted on 2-3 studies in the review. Pairs of authors [RM, SS, ZM, AS, LA, EK, ET] will independently extract the following data from included studies, resolve discrepancies and refer any unresolved to a third author [LA, RM]:

1. Methods: study authors, location, nature of service, period and length of study, period of comparator/s, disease (if applicable), and whether the changes in utilised services were likely due to them being omitted, delayed (or unclear).
2. Primary Outcome(s): percentage change in utilisation of health services and 95% CI, in pre and pandemic periods, and changes in absolute numbers of utilization, where data allow for calculation of percentage of change and 95% CI. In relation to the earlier point about packages of care, including care which flows from an initial indication or admission, when the data permits, we will consider the initial indication for the healthcare services utilisation as our primary outcome.

3. Secondary Outcome(s): change in the nature/characteristics of the users of health services (e.g. disease severity; disease spectrum/mix, or diagnostic yield; admissions to acute care)

#### Assessment of risk of bias in included studies

Pairs of review authors [RM, SS, ZM, AS, LA, EK, ET] will independently assess the risk of bias for each included study. We will use a modification of two risk of bias tools designed to assess before-after studies and interrupted time series analyses, the ROBINS-I tool [18-19] and a tool developed by the Cochrane EPOC group. [20] All disagreements will be resolved by discussion or by referring to a third author [RM, LA, AS, SS]. The following domains will be assessed:

1. Bias due to confounding (extraneous events)
2. Bias due to confounding (pre-intervention trends)
3. Bias in selection of participants
4. Bias due to missing data
5. Bias in measurement of the outcome
6. Bias in selection of reported result

Each potential source of bias will be graded as low, high or unclear, and each judgement was supported by a quote from the relevant trial. If secondary review outcomes require specific assessment on risk of bias domains this will be identified during further testing of the tool. Assessments of risk of bias will be presented for individual studies and across studies and will be incorporated into the results of the systematic review.

#### Data synthesis

We anticipate a wide heterogeneity in the population, settings, outcome measures, and methods used in the included studies, such that we do not expect to be able to perform a formal quantitative synthesis, i.e. a meta-analysis. Therefore, we plan to summarise the results narratively by using descriptive statistics, graphical figures, and a narrative synthesis. We will summarise the findings of included studies for the primary outcome grouped by service types: e.g. visits/admissions/consultations; diagnostic investigations; therapeutic/preventive interventions. If further sub-categorisation is needed, it will be by service locations: e.g. emergency department; primary care; and/or service specialty e.g. cardiology. We will calculate the mean difference and 95% confidence intervals for the change in the primary outcomes for each included study as appropriate.

If there is a sufficient number of sufficiently similar studies with acceptable levels of heterogeneity, and the data enable it, we would then aim to conduct a meta-analysis. In that case, we will use a random-effects model as the default to incorporate the assumption of heterogeneity between studies. We will evaluate statistical heterogeneity using both $Chi^2$ test (i.e. P value less than 0.10 was considered to be statistically significant heterogeneity) and the $I^2$ statistic (i.e. $I^2$ value of 0-40% was considered to be low heterogeneity, 40-60% moderate heterogeneity, 60-90% substantial heterogeneity, over 90% to be considerable heterogeneity).[19]

We anticipate that reporting of the secondary outcomes in each of the included studies will likely be expressed in a multitude of ways, specific to each study setting, disease category, patient population and category of utilisation. However, we will aim, if possible, to develop different categories for reporting of secondary outcomes.

#### Data Management

We will manage data using Endnote files, word documents and excel spreadsheets.

#### Dealing with missing data

If any primary studies only include changes as proportions, but do not include changes in absolute numbers of services, we will contact investigators or study sponsors to provide missing data.

#### Subgroup and sensitivity analyses

If there is a sufficient number of sufficiently similar studies with acceptable levels of heterogeneity to quantitatively synthesise the results, and the data enable it, we aim to conduct a sensitivity analysis (i) including only studies at an overall low risk of bias (eg low risk of bias in at least four of the six domains or interrupted time series studies vs pre-post pandemic studies); and (ii) including studies of longer duration (eg >6 weeks).

#### Assessment of reporting or publication biases

We plan to consider the possibility of the presence of reporting and/or publication bias and will take into account its likely influence when interpreting the review findings. If ten or more studies are included in a meta-analysis, we plan to examine the possibility of publication or small study bias using funnel plots. [19]

#### Additional analyses

We considered a range of analyses to explore correlations between study outcomes and other potentially relevant variables available outside the study data, such as nation-specific data about the stage of lockdown in the host nation at the time of the primary study. However, due to complexities in the large number of variables and potential discrepancies between official policy on restrictions and actual behaviour of people, as well as complex variation in the behaviours of different entities within the healthcare systems across the world, we decided, at protocol stage, to restrict our analysis to data within the publications.

#### Registration

We will register this protocol in the Open Science Framework, and in Prospero.

#### Sources of Support

The first author RM is funded by a National Health and Medical Research Council, NHMRC fellowship grant No 1124207 and is a chief investigator on an NHMRC Centre for Research Excellence, grant No 1104136. MJ is funded by The Foundation for Education and Development in Swedish Healthcare. AMS's salary is funded by the NHMRC CREMARC grant GNT 1153299. SS's position is supported by an NHMRC program grant. LA's salary is supported by an NHMRC CRE grant. The work does not necessarily represent the views of the organisations with which the authors are affiliated, or the funding bodies.

August 11, 2020

20. Cochrane Effective Practice and Organisation of Care (EPOC). Suggested risk of bias criteria for EPOC reviews. EPOC Resources for review authors 2017. Available at: Epoc.cochrane.org/resources/epoc-resources-review-authors

#### APPENDIX 1 – DATABASE SEARCH STRINGS

**PubMed**
("COVID-19"[Supplementary Concept] OR “COVID-19”[tiab] OR COVID19[tiab] OR “COVID 19”[tiab] OR “SARS-CoV-2”[tiab] OR “2019-nCoV”[tiab] OR “Novel coronavirus”[tiab] OR “Coronavirus 2019”[tiab] OR “Coronavirus 19”[tiab] OR “COVID 2019”[tiab] OR "2019 ncov"[tiab] OR “Wuhan coronavirus”[tiab])
AND
(((Pandemic[ti] OR Pandemics[ti] OR Outbreak[ti] OR Outbreaks[ti] OR Hospital[ti] OR Hospitals[ti] OR Emergency[ti] OR Surgery[ti] OR Surgical[ti] OR Department[ti] OR Departments[ti] OR Unit[ti] OR Units[ti] OR Clinic[ti] OR Clinics[ti] OR “Primary care”[ti])
AND
(Admission[ti] OR Admissions[ti] OR Visit[ti] OR Visits[ti] OR Attendance[ti] OR Attending[ti] OR Activity[ti] OR Utilization[ti] OR Utilisation[ti] OR Impact[ti] OR Impacts[ti] OR Reduction[ti] OR Reductions[ti] OR Decrease[ti] OR Decreases[ti] OR Decreased[ti] OR Decline[ti] OR Decline[ti] OR Change[ti] OR Changes[ti] OR Increase[ti] OR Increases[ti] OR Increased[ti]))
OR
((Pandemic[tiab] OR Pandemics[tiab] OR Outbreak[tiab] OR Outbreaks[tiab])
AND
(((Hospital[tiab] OR Hospitals[tiab] OR Emergency[tiab] OR Surgery[tiab] OR Surgical[tiab] OR Department[tiab] OR Departments[tiab] OR Unit[tiab] OR Units[tiab] OR Clinic[tiab] OR Clinics[tiab] OR “Primary care”[tiab] OR Telemedicine[tiab] OR Telehealth[tiab])
AND
(Admission[tiab] OR Admissions[tiab] OR Visit[tiab] OR Visits[tiab] OR Attendance[tiab] OR Attending[tiab] OR Activity[tiab] OR Utilization[tiab] OR Utilisation[tiab]))
OR
(Prescriptions[tiab] OR Prescribed[tiab] OR Vaccinations[tiab] OR Imaging[tiab] OR Scans[tiab] OR Endoscopy[tiab] OR Endoscopic[tiab] OR Endoscopies[tiab]))
AND
(Impact[tiab] OR Impacts[tiab] OR Reduction[tiab] OR Reductions[tiab] OR Decrease[tiab] OR Decreases[tiab] OR Decreased[tiab] OR Decline[tiab] OR Declines[tiab] OR Changes[tiab] OR Increase[tiab] OR Increases[tiab] OR Increased[tiab])))

**Embase (via Elsevier)**
(‘coronavirus disease 2019’/exp OR COVID-19:ti,ab OR COVID19:ti,ab OR "COVID 19":ti,ab OR SARS-CoV-2:ti,ab OR 2019-nCoV:ti,ab OR "Novel coronavirus":ti,ab OR "Coronavirus 2019":ti,ab OR "Coronavirus 19":ti,ab OR "COVID 2019":ti,ab OR "2019 ncov":ti,ab OR "Wuhan coronavirus":ti,ab)
AND
(((Pandemic:ti OR Pandemics:ti OR Outbreak:ti OR Outbreaks:ti OR Hospital:ti OR Hospitals:ti OR Emergency:ti OR Surgery:ti OR Surgical:ti OR Department:ti OR Departments:ti OR Unit:ti OR Units:ti OR Clinic:ti OR Clinics:ti OR "Primary care":ti)
AND
(Admission:ti OR Admissions:ti OR Visit:ti OR Visits:ti OR Attendance:ti OR Attending:ti OR Activity:ti OR Utilization:ti OR Utilisation:ti OR Impact:ti OR Impacts:ti OR Reduction:ti OR Reductions:ti OR Decrease:ti OR Decreases:ti OR Decreased:ti OR Decline:ti OR Decline:ti OR Change:ti OR Changes:ti

OR Increase:ti OR Increases:ti OR Increased:ti))
OR
((Pandemic:ti,ab OR Pandemics:ti,ab OR Outbreak:ti,ab OR Outbreaks:ti,ab)
AND
(((Hospital:ti,ab OR Hospitals:ti,ab OR Emergency:ti,ab OR Surgery:ti,ab OR Surgical:ti,ab OR Department:ti,ab OR Departments:ti,ab OR Unit:ti,ab OR Units:ti,ab OR Clinic:ti,ab OR Clinics:ti,ab OR "Primary care":ti,ab OR Telemedicine:ti,ab OR Telehealth:ti,ab)
AND
(Admission:ti,ab OR Admissions:ti,ab OR Visit:ti,ab OR Visits:ti,ab OR Attendance:ti,ab OR Attending:ti,ab OR Activity:ti,ab OR Utilization:ti,ab OR Utilisation:ti,ab))
OR
(Prescriptions:ti,ab OR Prescribed:ti,ab OR Vaccinations:ti,ab OR Imaging:ti,ab OR Scans:ti,ab OR Endoscopy:ti,ab OR Endoscopic:ti,ab OR Endoscopies:ti,ab))
AND
(Impact:ti,ab OR Impacts:ti,ab OR Reduction:ti,ab OR Reductions:ti,ab OR Decrease:ti,ab OR Decreases:ti,ab OR Decreased:ti,ab OR Decline:ti,ab OR Declines:ti,ab OR Changes:ti,ab OR Increase:ti,ab OR Increases:ti,ab OR Increased:ti,ab)))

**Cochrane COVID-19 Study Register**
Pandemic OR Pandemics OR Outbreak OR Outbreaks
AND
(Hospital OR Hospitals OR Emergency OR Surgery OR Surgical OR Department OR Departments OR Unit OR Units OR Clinic OR Clinics OR "Primary care" OR Telemedicine OR Telehealth)
AND
(Admission OR Admissions OR Visit OR Visits OR Attendance OR Attending OR Activity OR Utilization OR Utilisation OR Prescriptions OR Prescribed OR Vaccinations OR Imaging OR Scans OR Endoscopy OR Endoscopic OR Endoscopies)
AND
(Impact OR Impacts OR Reduction OR Reductions OR Decrease OR Decreases OR Decreased OR Decline OR Declines OR Changes OR Increase OR Increases OR Increased)

**Europe PMC preprints**
(COVID-19 OR COVID19 OR "COVID 19" OR SARS-CoV-2 OR 2019-nCoV OR "Novel coronavirus" OR "Coronavirus 2019" OR "Coronavirus 19" OR "COVID 2019" OR "2019 ncov" OR "Wuhan coronavirus")
AND
(Pandemic[ti] OR Pandemics[ti] OR Outbreak[ti] OR Outbreaks[ti])
AND
(Hospital OR Hospitals OR Emergency OR Surgery OR Surgical OR Department OR Departments OR Unit OR Units OR Clinic OR Clinics OR "Primary care" OR Telemedicine OR Telehealth)
AND
(Admission OR Admissions OR Visit OR Visits OR Attendance OR Attending OR Activity OR Utilization OR Utilisation OR Prescriptions OR Prescribed OR Vaccinations OR Imaging OR Scans OR Endoscopy OR Endoscopic OR Endoscopies)
AND
(Impact OR Impacts OR Reduction OR Reductions OR Decrease OR Decreases OR Decreased OR Decline OR Declines OR Changes OR Increase OR Increases OR Increased)

#### Supplementary file 2

Checklist of items to include when reporting a systematic review or meta-analysis

| Section/topic | # | Checklist item | Reported on page # |
|---|---|---|---|
| **TITLE** | | | |
| Title | 1 | Identify the report as a systematic review, meta-analysis, or both. | 1 |
| **ABSTRACT** | | | |
| Structured summary | 2 | Provide a structured summary including, as applicable: background; objectives; data sources; study eligibility criteria, participants, and interventions; study appraisal and synthesis methods; results; limitations; conclusions and implications of key findings; systematic review registration number. | 3-4 |
| **INTRODUCTION** | | | |
| Rationale | 3 | Describe the rationale for the review in the context of what is already known. | 5 |
| Objectives | 4 | Provide an explicit statement of questions being addressed with reference to participants, interventions, comparisons, outcomes, and study design (PICOS). | 5 |
| **METHODS** | | | |
| Protocol and registration | 5 | Indicate if a review protocol exists, if and where it can be accessed (e.g., Web address), and, if available, provide registration information including registration number. | 6 |
| Eligibility criteria | 6 | Specify study characteristics (e.g., PICOS, length of follow-up) and report characteristics (e.g., years considered, language, publication status) used as criteria for eligibility, giving rationale. | 6 |
| Information sources | 7 | Describe all information sources (e.g., databases with dates of coverage, contact with study authors to identify additional studies) in the search and date last searched. | 6,7 |
| Search | 8 | Present full electronic search strategy for at least one database, including any limits used, such that it could be repeated. | Supp. file 3 |

| Section/topic | # | Checklist item | Reported on page # |
|---|---|---|---|
| Study selection | 9 | State the process for selecting studies (i.e., screening, eligibility, included in systematic review, and, if applicable, included in the meta-analysis). | 6,7 |
| Data collection process | 10 | Describe method of data extraction from reports (e.g., piloted forms, independently, in duplicate) and any processes for obtaining and confirming data from investigators. | 7,8 |
| Data items | 11 | List and define all variables for which data were sought (e.g., PICOS, funding sources) and any assumptions and simplifications made. | 8 |
| Risk of bias in individual studies | 12 | Describe methods used for assessing risk of bias of individual studies (including specification of whether this was done at the study or outcome level), and how this information is to be used in any data synthesis. | 7 |
| Summary measures | 13 | State the principal summary measures (e.g., risk ratio, difference in means). | 8 |
| Synthesis of results | 14 | Describe the methods of handling data and combining results of studies, if done, including measures of consistency (e.g., $I^2$) for each meta-analysis. | 8 |
| Risk of bias across studies | 15 | Specify any assessment of risk of bias that may affect the cumulative evidence (e.g., publication bias, selective reporting within studies). | 7 |
| Additional analyses | 16 | Describe methods of additional analyses (e.g., sensitivity or subgroup analyses, meta-regression), if done, indicating which were pre-specified. | 8 |
| **RESULTS** | | | |
| Study selection | 17 | Give numbers of studies screened, assessed for eligibility, and included in the review, with reasons for exclusions at each stage, ideally with a flow diagram. | 9 |
| Study characteristics | 18 | For each study, present characteristics for which data were extracted (e.g., study size, PICOS, follow-up period) and provide the citations. | 9 |
| Risk of bias within studies | 19 | Present data on risk of bias of each study and, if available, any outcome-level assessment (see Item 12). | 9,10 |
| Results of individual studies | 20 | For all outcomes considered (benefits or harms), present, for each study: (a) simple summary data for each intervention group and (b) effect estimates and | 10-12 |

| Section/topic | # | Checklist item | Reported on page # |
|---|---|---|---|
| | | confidence intervals, ideally with a forest plot. | |
| Synthesis of results | 21 | Present results of each meta-analysis done, including confidence intervals and measures of consistency. | N/A |
| Risk of bias across studies | 22 | Present results of any assessment of risk of bias across studies (see Item 15). | Figure 2, and Supp. File 4 |
| Additional analysis | 23 | Give results of additional analyses, if done (e.g., sensitivity or subgroup analyses, meta-regression) (see Item 16). | N/A |
| **DISCUSSION** | | | |
| Summary of evidence | 24 | Summarize the main findings including the strength of evidence for each main outcome; consider their relevance to key groups (e.g., health care providers, users, and policy makers). | 12 |
| Limitations | 25 | Discuss limitations at study and outcome level (e.g., risk of bias), and at review level (e.g., incomplete retrieval of identified research, reporting bias). | 12,13 |
| Conclusions | 26 | Provide a general interpretation of the results in the context of other evidence, and implications for future research. | 13, 14 |
| **FUNDING** | | | |
| Funding | 27 | Describe sources of funding for the systematic review and other support (e.g., supply of data); role of funders for the systematic review. | Abstract |

#### Supplementary File 3 – DATABASE SEARCH STRINGS

**PubMed**

("COVID-19"[Supplementary Concept] OR “COVID-19”[tiab] OR COVID19[tiab] OR “COVID 19”[tiab] OR “SARS-CoV-2”[tiab] OR “2019-nCoV”[tiab] OR “Novel coronavirus”[tiab] OR “Coronavirus 2019”[tiab] OR “Coronavirus 19”[tiab] OR “COVID 2019”[tiab] OR "2019 ncov"[tiab] OR “Wuhan coronavirus”[tiab])
AND
(((Pandemic[ti] OR Pandemics[ti] OR Outbreak[ti] OR Outbreaks[ti] OR Hospital[ti] OR Hospitals[ti] OR Emergency[ti] OR Surgery[ti] OR Surgical[ti] OR Department[ti] OR Departments[ti] OR Unit[ti] OR Units[ti] OR Clinic[ti] OR Clinics[ti] OR “Primary care”[ti])
AND
(Admission[ti] OR Admissions[ti] OR Visit[ti] OR Visits[ti] OR Attendance[ti] OR Attending[ti] OR Activity[ti] OR Utilization[ti] OR Utilisation[ti] OR Impact[ti] OR Impacts[ti] OR Reduction[ti] OR Reductions[ti] OR Decrease[ti] OR Decreases[ti] OR Decreased[ti] OR Decline[ti] OR Decline[ti] OR Change[ti] OR Changes[ti] OR Increase[ti] OR Increases[ti] OR Increased[ti]))
OR
((Pandemic[tiab] OR Pandemics[tiab] OR Outbreak[tiab] OR Outbreaks[tiab])
AND
(((Hospital[tiab] OR Hospitals[tiab] OR Emergency[tiab] OR Surgery[tiab] OR Surgical[tiab] OR Department[tiab] OR Departments[tiab] OR Unit[tiab] OR Units[tiab] OR Clinic[tiab] OR Clinics[tiab] OR “Primary care”[tiab] OR Telemedicine[tiab] OR Telehealth[tiab])
AND
(Admission[tiab] OR Admissions[tiab] OR Visit[tiab] OR Visits[tiab] OR Attendance[tiab] OR Attending[tiab] OR Activity[tiab] OR Utilization[tiab] OR Utilisation[tiab]))
OR
(Prescriptions[tiab] OR Prescribed[tiab] OR Vaccinations[tiab] OR Imaging[tiab] OR Scans[tiab] OR Endoscopy[tiab] OR Endoscopic[tiab] OR Endoscopies[tiab]))
AND
(Impact[tiab] OR Impacts[tiab] OR Reduction[tiab] OR Reductions[tiab] OR Decrease[tiab] OR Decreases[tiab] OR Decreased[tiab] OR Decline[tiab] OR Declines[tiab] OR Changes[tiab] OR Increase[tiab] OR Increases[tiab] OR Increased[tiab])))

**Embase (via Elsevier)**

(‘coronavirus disease 2019’/exp OR COVID-19:ti,ab OR COVID19:ti,ab OR "COVID 19":ti,ab OR SARS-CoV-2:ti,ab OR 2019-nCoV:ti,ab OR "Novel coronavirus":ti,ab OR "Coronavirus 2019":ti,ab OR "Coronavirus 19":ti,ab OR "COVID 2019":ti,ab OR "2019 ncov":ti,ab OR "Wuhan coronavirus":ti,ab)
AND
(((Pandemic:ti OR Pandemics:ti OR Outbreak:ti OR Outbreaks:ti OR Hospital:ti OR Hospitals:ti OR Emergency:ti OR Surgery:ti OR Surgical:ti OR Department:ti OR Departments:ti OR Unit:ti OR Units:ti OR Clinic:ti OR Clinics:ti OR "Primary care":ti)
AND
(Admission:ti OR Admissions:ti OR Visit:ti OR Visits:ti OR Attendance:ti OR Attending:ti OR Activity:ti OR Utilization:ti OR Utilisation:ti OR Impact:ti OR Impacts:ti OR Reduction:ti OR Reductions:ti OR Decrease:ti OR Decreases:ti OR Decreased:ti OR Decline:ti OR Decline:ti OR Change:ti OR Changes:ti OR Increase:ti OR Increases:ti OR Increased:ti))
OR
((Pandemic:ti,ab OR Pandemics:ti,ab OR Outbreak:ti,ab OR Outbreaks:ti,ab)
AND
(((Hospital:ti,ab OR Hospitals:ti,ab OR Emergency:ti,ab OR Surgery:ti,ab OR Surgical:ti,ab OR

Department:ti,ab OR Departments:ti,ab OR Unit:ti,ab OR Units:ti,ab OR Clinic:ti,ab OR Clinics:ti,ab OR "Primary care":ti,ab OR Telemedicine:ti,ab OR Telehealth:ti,ab)
AND
(Admission:ti,ab OR Admissions:ti,ab OR Visit:ti,ab OR Visits:ti,ab OR Attendance:ti,ab OR Attending:ti,ab OR Activity:ti,ab OR Utilization:ti,ab OR Utilisation:ti,ab))
OR
(Prescriptions:ti,ab OR Prescribed:ti,ab OR Vaccinations:ti,ab OR Imaging:ti,ab OR Scans:ti,ab OR Endoscopy:ti,ab OR Endoscopic:ti,ab OR Endoscopies:ti,ab))
AND
(Impact:ti,ab OR Impacts:ti,ab OR Reduction:ti,ab OR Reductions:ti,ab OR Decrease:ti,ab OR Decreases:ti,ab OR Decreased:ti,ab OR Decline:ti,ab OR Declines:ti,ab OR Changes:ti,ab OR Increase:ti,ab OR Increases:ti,ab OR Increased:ti,ab)))

**Cochrane COVID-19 Study Register**
Pandemic OR Pandemics OR Outbreak OR Outbreaks
AND
(Hospital OR Hospitals OR Emergency OR Surgery OR Surgical OR Department OR Departments OR Unit OR Units OR Clinic OR Clinics OR "Primary care" OR Telemedicine OR Telehealth)
AND
(Admission OR Admissions OR Visit OR Visits OR Attendance OR Attending OR Activity OR Utilization OR Utilisation OR Prescriptions OR Prescribed OR Vaccinations OR Imaging OR Scans OR Endoscopy OR Endoscopic OR Endoscopies)
AND
(Impact OR Impacts OR Reduction OR Reductions OR Decrease OR Decreases OR Decreased OR Decline OR Declines OR Changes OR Increase OR Increases OR Increased)

**Europe PMC preprints**
(COVID-19 OR COVID19 OR "COVID 19" OR SARS-CoV-2 OR 2019-nCoV OR "Novel coronavirus" OR "Coronavirus 2019" OR "Coronavirus 19" OR "COVID 2019" OR "2019 ncov" OR "Wuhan coronavirus")
AND
(Pandemic[ti] OR Pandemics[ti] OR Outbreak[ti] OR Outbreaks[ti])
AND
(Hospital OR Hospitals OR Emergency OR Surgery OR Surgical OR Department OR Departments OR Unit OR Units OR Clinic OR Clinics OR "Primary care" OR Telemedicine OR Telehealth)
AND
(Admission OR Admissions OR Visit OR Visits OR Attendance OR Attending OR Activity OR Utilization OR Utilisation OR Prescriptions OR Prescribed OR Vaccinations OR Imaging OR Scans OR Endoscopy OR Endoscopic OR Endoscopies)
AND
(Impact OR Impacts OR Reduction OR Reductions OR Decrease OR Decreases OR Decreased OR Decline OR Declines OR Changes OR Increase OR Increases OR Increased)

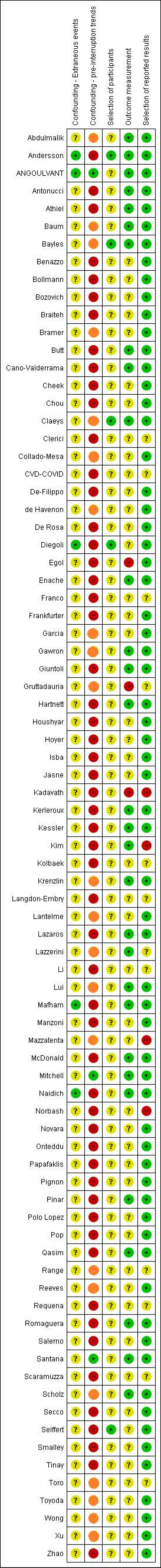
Confounding - Extraneous events
Confounding - pre-interruption trends
Selection of participants
Outcome measurement
Selection of reported results
Abdulmalik
Andersson
ANGOULVANT
Antonucci
Athiel
Baum
Bayles
Benazzo
Bollmann
Bozovich
Braiteh
Bramer
Butt
Cano-Valderrama
Cheek
Chou
Claeys
Clerici
Collado-Mesa
CVD-COVID
De-Filippo
de Havenon
De Rosa
Diegoli
Egol
Enache
Franco
Frankfurter
Garcia
Gawron
Giuntoli
Gruttadauria
Hartnett
Houshyar
Hoyer
Isba
Jasne
Kadavath
Kerleroux
Kessler
Kim
Kolbaek
Krenzlin
Langdon-Embry
Lantelme
Lazaros
Lazzerini
Li
Lui
Mafham
Manzoni
Mazzatenta
McDonald
Mitchell
Naidich
Norbash
Novara
Onteddu
Papafaklis
Pignon
Pinar
Polo Lopez
Pop
Qasim
Range
Reeves
Requena
Romaguera
Salerno
Santana
Scaramuzza
Scholz
Secco
Seiffert
Smalley
Tinay
Toro
Toyoda
Wong
Xu
Zhao

### Title: Pandemic impacts on healthcare utilisation: a systematic review

Authors names:  R Moynihan, S Sanders, ZA Michaleff, AM Scott, J Clark, EJ To, M Jones, E Kitchener, M Fox, M Johansson, E Lang, A Duggan, IA Scott, L Albarqouni.

**Supplementary File 5** –

5.1 Table of Study Characteristics and reference list of all included studies.

5.2 Table of Results of the primary outcome of the included studies

5.3 Table of Results of secondary outcomes of the included studies

5.4 Figures of changes in healthcare utilisations reported in included studies

#### Supplementary Table. Characteristics of Included Studies of pandemic related changes in healthcare utilization

| Author; Country; Scope; Design | Setting; Population | Pandemic and comparator periods* | Primary Outcomes | Secondary Outcomes |
|---|---|---|---|---|
| Abdulmalik; Qatar; National; Same period single year | Outpatient/Primary care; 27 primary health care centres | March - May; 2020 vs. 2018-19 | Overall utilization of all primary healthcare services across all health centres | N/A |
| Andersson; Denmark; National; Same period single year | Hospital; Danish Nationwide Patient Registry | March 12 - March 31; 2020 vs. 2019 | Incidence rates of new-onset HF and hospitalization for worsening HF | Mortality |
| Angoulvant; France; Multi-centre; Time trend multiple years | ED & Hospital; 6 Paediatric EDs from academic hospitals being part of Assistance Publique – Hôpitaux de Paris | March 18 - April 19; 2020 vs. 2017-19 | Number of hospital visits and admissions | N/A |
| Antonucci; Italy; Multi-centre; Same period single year | ED & Hospital; 3 high volume urology departments in Rome, Italy | March - April; 2020 vs. 2019 | Number of ED admissions for urolithiasis; Number of hospitalisations | N/A |
| Athiel; France; Multi-centre; Same period single year | ED & Hospital; 12 gynaecological emergency units of the Greater Paris University Hospitals | March - May; 2020 vs. 2019 | Number of emergency gynaecological hospitalisations | N/A |

| | | | | |
|---|---|---|---|---|
| Baum; USA; National; Time trend single year | Hospital; Veterans Affairs Hospitals' Corporate Data Warehouse, a national repository of electronic health records from visits to any VA facility | March 11 – April 21; 2020 vs 2019 | All admissions for any condition | N/A |
| Bayles; USA; Multi-centre; Same period single year | ED; 3 acute care facilities from the Marin County Department of Health and Human Services | March 17 - May 4; 2020 vs. 2018-19 | Average number of daily ED visits | N/A |
| Benazzo; Italy; Multi-centre; Same period single year | ED & Hospital; 15 orthopaedic and trauma units | February 23 - April 4; 2020 vs. 2019 | Outpatient consultations; Trauma ED visits; Surgeries | N/A |
| Bollman; Germany; Multi-centre; Same period single year | Hospital; 66 Helios hospitals | March 1 - April 30; 2020 vs. 2019 | Admissions for heart failures and arrythmias | N/A |
| Bozovich; Argentina; Multi-centre; Same period single year | ED & Hospital; 31 private hospitals | April 1 - April 30; 2020 vs. 2019 | ED consultations and procedures | N/A |
| Braiteh; USA; Multi-centre; Same period single year | Hospital; 4 hospitals | March - April; 2020 vs. 2019 | Admissions for any cause; Presentations for Acute Coronary Syndrome (also describes as admissions) | Rates of STEMI versus NSTEMI |

| | | | | |
|---|---|---|---|---|
| Bramer; USA; Multi-centre; Same period single year | Community; vaccinations from one state immunization system | May; 2020 vs. 2017-19 | Proportion of children with up-to-date status for all recommended vaccines | N/A |
| Butt; Qatar; Multi-centre; Same period single year | ED; 2 hospitals in Qatar that see over 80% of patients in Qatar with suspected Acute Coronary Syndrome | March - April; 2020 vs. 2019 | Total ED visits; ED presentations with cardiac symptoms | Rates presenting with Acute Coronary Syndrome (ACS) |
| Cano-Valderrama; Spain; Multi-centre; Same period single year | Hospital; 3 tertiary care centres | March 16 - April 26; 2020 vs. 2019 | Acute care surgeries | SOFA scores |
| Cheek; Australia; Multi-centre; Same period single year | ED; 2 tertiary hospitals and 2 urban district hospitals | March 22 - May 23; 2020 vs. 2019 | Number of attendances at paediatric ED; Number of attendances at paediatric ED for mental health diagnoses; Number of neonatal presentations | N/A |
| Chou; Taiwan; Multi-centre; Same period single year | Community/Primary care; Hospice homecare services, hospice inpatient services and non-hospice services provided by 2 branches of health care organisation in Northern Taiwan | January - April; 2020 vs. 2019 | Number of hospice home care visits; Number of new enrolments in hospice home care; Bed occupancy rates in hospice and non-hospice units; Monthly patient days in hospice and non-hospice units | N/A |
| Claeys; Belgium; National; Same period single year | Hospital; 36 of the 49 PCI-capable hospitals in the Belgian STEMI database and Belgian Coronary Stent Registry | March 13 - April 3; 2020 vs. 2017-19 | Number of STEMI admission | Mortality; % cardiac arrest; Killip class |

| | | | | |
|---|---|---|---|---|
| Clerici; Italy; Multi-centre; Same period single year | Hospital; 7 general hospital psychiatric wards in the Lombardy region of Italy | February 21 - March 31; 2020 vs. 2019 | Average daily number of admissions by week, total number of weekly admissions; Annual rates of admissions/1000 adults | Number of voluntary and involuntary admissions |
| Collado-Mesa; USA; Multi-centre; Same period single year | Community/Outpatient; five breast imaging centres | April; 2020 vs. 2018-19 | Number of breast imaging examinations; Number of image-guided procedures | Proportion of positive biopsy of image guided biopsy |
| CVD-Covid-UK Consortium; UK; Multi-centre; Same period single year | Hospital; 9 hospitals in England and Scotland | March 23 - May 10; 2020 vs. 2018-19 | Number of ED attendances and hospital admissions | procedures for cardiac, cerebrovascular, other vascular conditions |
| De Filippo; Italy; Multi-centre; Same period single year | Hospital; 15 hospitals in Northern Italy | February 20 - March 31; 2020 vs. 2019 | Incidence rate ratio for hospital admissions for ACS | Incidence rate ratio for STEMI/NSTEMI |
| de Havenon; USA; Multi-centre; Same period single year | Hospital; 65 academic and community hospitals | February - March; 2020 vs. 2018-19 | Number of hospitalisations for stroke and ACS; Number of procedures for stroke and ACS | N/A |
| De Rosa; Italy; Multi-centre; Same period single year | Hospital; cardiac care units at 54 Italian hospitals affiliated with Italian Society of Cardiology | March 12 - March 19; 2020 vs. 2019 | Number of admissions for acute myocardial infarction | Case fatality rates; Number of admissions per diagnosis (STEMI/NSTEMI) |

| | | | | |
|---|---|---|---|---|
| Diegoli; Brazil; Multi-centre; Same period single year | Hospital; 6 hospitals in Joinville, Brazil | March 17 - April 15; 2020 vs. 2019 | Admissions for stroke/100000 inhabitants | Admissions for severe stroke (NIH stroke scale score) |
| Egol; USA; Multi-centre; Same period single year | ED & Hospital; The NYU Langone Orthopaedic Department is responsible for the musculoskeletal care at 7 different hospitals within the New York City area. | February 1 - April 15; 2020 vs. 2019 | Number of ED presentations with hip fracture | Mortality; Non/operative case |
| Enache; Monaco; National; Same period single year | ED & Hospital; Monaco public health care system | March; 2020 vs. 2019 | Number cardiovascular and emergency admissions | N/A |
| Franco; Italy; Multi-centre; Same period single year | Hospital; 10 cardiology centres in Northern Italy | February 23 - March 28; 2020 vs. 2019 | Number of hospitalisations for NSTEMI | N/A |
| Frankfurter; Canada; Multi-centre; Same period single year | ED & Hospital; University Health Network (Toronto General Hospital and Toronto Western Hospital), in Toronto, Canada | March 1 - April 19; 2020 vs. 2019 | Number ED visits and hospitalised with heart failure | ICU admission; Mortality; Hospitalisation; NYHA class III-IV |
| Garcia; USA; Multi-centre; Time trend single year | Hospital; 18 sites representing primary percutaneous coronary intervention (PPCI) hospitals and healthcare systems across the US | March - April; 2020 vs. 2019 | Monthly volume of cardiac catheterisation leading to intervention (angiography) | N/A |

| | | | | |
|---|---|---|---|---|
| Gawron; USA; National; Time trend single year | Hospital & Outpatient; 170 medical centres and 1074 outpatient sites | March - April; 2020 vs. 2019 | Average number of upper gastrointestinal endoscopies per month | N/A |
| Giuntoli; Italy; Multi-centre; Same period single year | Hospital; three of the major trauma and elective orthopaedic surgery centres of north-west Tuscany | March; 2020 vs. 2019 | Number of patients treated | Hospitalisation |
| Gruttadauria; Italy; Multi-centre; Same period single year | Hospital; 22 Italian Liver Transplant Programs. | March 1 - March 15; 2020 vs. 2018-19 | Number of liver transplants | N/A |
| Hartnett; USA; Multi-centre; Same period single year | ED; subset of hospitals in 47 states capturing approximately 73% of ED visits in the USA | March 29 - April 25; 2020 vs. 2019 | Mean weekly ED presentations | N/A |
| Houshyar; USA; Multi-centre; Same period single year | ED & Hospital; 5 University of California Health Centres with academic radiology programs. | March 19 - April 2; 2020 vs. 2019 | Daily number of ED radiologic examinations | N/A |
| Hoyer; Germany; Multi-centre; Same period single year | ED & Hospital; 4 German comprehensive stroke centres. | March 16 - April 12; 2020 vs. 2019 | Numbers of patients admitted with final diagnoses of ischemic stroke or TIA | TIA/ Stroke |

| | | | | |
|---|---|---|---|---|
| Isba; UK; Multi-centre; Same period single year | ED; 2 hospitals in greater Manchester | February - March; 2020 vs. 2019 | Weekly PED attendances | N/A |
| Jasne; USA; Multi-centre; Same period single year | ED & Hospital; 3 hospitals in New Haven, Connecticut | March 1 - April 28; 2020 vs. 2019 | Weekly stoke code calls | N/A |
| Kadavath; USA; Multi-centre; Same period single year | Hospital; 12 fellowship training sties | March 1 - April 15; 2020 vs. 2019 | Number of invasive cardiac procedures | N/A |
| Kerleroux; France; Multi-centre; Same period single year | Hospital; 32 centres in all French administrative regions. | February 15 - March 30; 2020 vs. 2019 | Number of patients receiving MT between study periods | % unwitnessed onset; Baseline NIHSS; ASPECTs |
| Kessler; Germany; Multi-centre; Same period single year | Hospital; 15 cardiac care centres distributed across Germany providing 24/7 interventional cardiac care. | March 1 - April 30; 2020 vs. 2019 | Number of patients presenting with Acute Coronary Syndrome | STEMI/NSTEMI |
| Kim; USA; Multi-centre; Same period single year | ED; seven EDs include one urban academic hospital, five suburban community hospitals, and one free-standing ED. | March 8 - May 2; 2020 vs. 2019 | Weekly Emergency Department visits | N/A |

| | | | | |
|---|---|---|---|---|
| Kolbaek; Denmark; Multi-centre; Same period single year | Community/Outpatient; Psychiatric services | February 23 - May 2; 2020 vs. 2019 | Number of referrals to psychiatric service | N/A |
| Krenzlin; Germany; Multi-centre; Same period single year | Hospital; Two major neurosurgical departments in Germany | March 16 - April 19; 2020 vs. 2018-19 | Number of emergency admissions | N/A |
| Langdon-Embry; USA; Multi-centre; Same period single year | Community; childhood immunisation facilities in New York City | March 16 – May 31; 2020 vs. 2019 | Number of childhood vaccine doses administered; Number of unique facilities reporting administration of at least one childhood vaccine | N/A |
| Lantelme; France; Multi-centre; Same period single year | Hospital; 3 public centres in Lyon. | March 9 - April 5; 2020 vs. 2019 | Weekly rate of hospital admissions for myocardial infarction | N/A |
| Lazaros; Greece; Multi-centre; Same period single year | Hospital; 2 large hospitals of the National Health System belonging to the larger Metropolitan area of Athens | March 12 - May 7; 2020 vs. 2019 | Number of cardiac surgery procedures | Emergency vs non-emergency |
| Lazzerini; Italy; Multi-centre; Same period single year | ED; 5 Pediatric ED (three third-level referral hospitals and two second-level hospitals) | March 1 - March 27; 2020 vs. 2019 | Number of paediatric emergency department visits | N/A |

| | | | | |
|---|---|---|---|---|
| Li; Taiwan; Multi-centre; Same period single year | Hospital; 40 major hospitals | February - April; 2020 vs. 2019 | Number of patients admitted for STEMI | N/A |
| Lui; Hong Kong; National; Same period single year | Hospital; all public hospitals | January 21 - March 31; 2020 vs. 2017-19 | Upper and lower endoscopies | Positive rate for colon cancer and gastric cancer |
| Mafham; UK; National; Same period single year | Hospital; 147 acute NHS hospital trusts | January 6 - May 30; 2020 vs. 2019 | Admissions for Acute Coronary Syndromes | Proportions of STEMI vs NSTEMI |
| Manzoni; Italy; Multi-centre; Same period single year | ED; 2 emergency paediatric departments | March - April; 2020 vs. 2019 | Volume of ED visits | Hospitalisation |
| Mazzatenta; Italy; Multi-centre; Same period single year | Hospital; 5 neurosurgery departments and 1 paediatric centre | March 13 - April 13; 2020 vs. 2018-19 | Outpatient consultations; Surgical activities | Urgent/nonurgent surgery |
| McDonald; UK; National; Same period single year | Community; electronic patient records of vaccination | March 2 - April 25; 2020 vs. 2019 | Hexavalent vaccines; MMR first vaccination | N/A |

| | | | | |
|---|---|---|---|---|
| Mitchell; Australia; Multi-centre; Time trend multiple years | ED & Hospital; 2 Emergency Departments | March 26 - April 25; 2020 vs. 2017-19 | Daily number of ED presentations | Triage category |
| Naidich; USA; Multi-centre; Same period single year | Hospital & Outpatient; 92 centres across NY state | March 2 - April 18; 2020 vs. 2019 | Volume of imaging | N/A |
| Norbash; USA; Multi-centre; Same period single year | Hospital & Outpatient; 6 academic medical systems | January 6 - May 23; 2020 vs. 2019 | Volume of imaging | N/A |
| Novara; Italy; Multi-centre; Same period single year | ED; EDs within 8 academic and non-academic urology centres | March 12 - March 16; 2020 vs. 2019 | ED urological consults | Triage category/ hospitalisation |
| Onteddu; Multi-national; Multi-centre; Same period single year | Hospital; TriNetX, a global health collaborative clinical research platform collecting real-time electronic medical record data from a network of health care organizations | January 20 - May 16; 2020 vs. 2019 | Number of ischemic stroke patients | N/A |
| Papafakis; Greece; Multi-centre; Same period single year | Hospital; Greek public hospitals with PCI capability, including a primary PCI service | March 2 - April 12; 2020 vs. 2019 | Number of patients admitted for Acute coronary syndrome | ACS presentation |

| | | | | |
|---|---|---|---|---|
| Pignon; France; Multi-centre; Same period single year | ED; 3 psychiatric emergency services | March 17 - April 13; 2020 vs. 2019 | Emergency psychiatric consultations | Rates of hospitalisation |
| Pinar; France; Multi-centre; Same period single year | Hospital; 8 academic urology departments | March 12 - March 27; 2020 vs. 2019 | Urological surgeries | N/A |
| Polo Lopez; Spain; Multi-centre; Same period single year | Hospital; 13 public hospitals where most congenital heart disease surgery in Spain is performed | March 13 - May 13; 2020 vs. 2019 | Number of congenital heart disease surgeries | N/A |
| Pop; France; Multi-centre; Same period single year | Hospital; 3 hospitals with stroke units | March 1 -March 31; 2020 vs. 2019 | Stroke alerts (following initial consult) | Proportion of alerts resulting in admissions for stroke; Initial NIHSS score |
| Qasim; USA; Multi-centre; Same period single year | ED; 4 adult and 2 paediatric Level 1 Trauma centres | March 9 - April 19; 2020 vs. 2019 | Trauma contacts | Rates of highest acuity (“alerts”) |
| Range; France; Multi-centre; Time trend single year | Hospital; 12 interventional cardiology centres | March 15 - April 4; 2020 vs. 2019 | Patients enrolled in Percutaneous Coronary Intervention registry (follows all STEMI patients undergoing PCI) | N/A |

| | | | | |
|---|---|---|---|---|
| Reeves; UK; Multi-centre; Time trend multiple years | Hospital; University hospitals in one NHS Foundation Trust | March 22 - April 25; 2020 vs. 2016-19 | Admissions for STEMI and stroke | N/A |
| Requena; Multi-national; Multi-centre; Same period single year | Community; 2 fertility facilities in Spain and 1 in Italy | February 3 - March 23; 2020 vs. 2019 | Fertility related procedures | N/A |
| Romaguera; Spain; Multi-centre; Same period single year | Hospital; 10 percutaneous coronary intervention hospitals | March 1 - April 19; 2020 vs. 2019 | STEMI admissions | Proportion of more severe Killip classes; Proportion of sudden cardiac death; mortality |
| Scaramuzza; Italy; Multi-centre; Same period single year | ED; 2 paediatric emergency departments | February 20 - March 30; 2020 vs. 2019 | Presentations to paediatric ED | Reductions across different triage categories |
| Salerno; Italy; National; Same period single year | Hospital; 35 endoscopy units in Italy | March; 2020 vs. 2019 | Number of urgent endoscopic procedures | Proportion of positive procedures (i.e. diagnostic yield) for urgent EGDs and lower endoscopy |
| Santana; Portugal; National; Time trend multiple years | ED; emergency services in mainland Portugal | March; 2020 vs. 2019 | Number of emergency episodes | Triage category |

| | | | | |
|---|---|---|---|---|
| Scholz; Germany; Multi-centre; Same period single year | Hospital; 41 percutaneous coronary intervention centres participating in a trial | March; 2020 vs. 2017-19 | Number of STEMI patients treated | Mortality; TIMI score |
| Secco; Italy; Multi-centre; Same period single year | Hospital; 3 high volume centres in North and Central Italy | March; 2020 vs. 2019 | Number of admissions for ACS | Type of ACS; TIMI score; GRACE score; Admission peak hs-troponin; Mortality |
| Seiffert; Germany; National; Same period single year | Hospital; Health insurance claims from second largest insurer in Germany | March 2 - May 31; 2020 vs. 2019 | Rate of admissions/100000 insured for cardiovascular or cerebrovascular emergencies | Number per diagnosis (STEMI, NSTEMI, stroke, TIA); Number of invasive procedures; Mortality |
| Smalley; USA; Multi-centre; Same period single year | ED; 20 EDs across a large Midwest integrated healthcare system | March 25 - April 24; 2020 vs. 2019 | Number of ED encounters; Number of behavioural health visits to the ED | N/A |
| Tinay; Turkey; Multi-centre; Same period single year | Hospital; Surgical urologic oncology practices | March 11-April 11; 2020 vs. 2019 | Number of nondeferrable uro-oncological procedures | ASA score |
| Toro; Chile; National; Time trend multiple years | ED; public health hospitals, emergency care services in 16 regions of Chile | March 8 - April 18; 2020 vs. 2015-19 | Number of emergency service consultations | N/A |

| | | | | |
|---|---|---|---|---|
| Toyoda; Multi-national; Multi-centre; Same period single year | Hospital; 3 liver speciality clinics | February 1 - May 1; 2020 vs. 2018-19 | Number of clinic visits; Number of ultrasounds performed; Number of CT/MRIs performed | Visits in advanced disease patients |
| Wong; Hong Kong; National; Same period single year | Hospital & Outpatient; 43 Hong Kong public hospitals and 122 outpatient clinics | January 25 - March 27; 2020 vs. 2016-19 | Mean weekly orthopaedic operations; Mean weekly orthopaedic emergencies treated operatively | Elective and emergency operations |
| Xu; USA; Multi-centre; Same period single year | Outpatient; retinal care centres | March 8 - May 16; 2020 vs. 2018-19 | Mean weekly office visits; Mean weekly intravitreal injections; Mean weekly optical coherence tomography, fluorescein angiography and indocyanine green testing | N/A |
| Zhao; China; Multi-centre; Same period single year | Hospital; 280 stroke centres across China participating in Big Data Observatory platform | January - February; 2020 vs. 2019 | Number of stroke admissions; Number of thrombolysis treatments; Number of thrombectomy treatments | N/A |

Abbreviations: CT: Computed Tomography Scan; ED: Emergency Department; MRI: Magnetic resonance imaging; N/A: Not applicable; NIHSS:NIH Stroke Scale Score; NSTEMI: Non-ST elevation myocardial infarction; PED: Paediatric Emergency Department; STEMI: ST-elevation myocardial infarction; TIA: Transient Ischaemic Attack.

Note: *This is the period of time analysed in this Systematic Review, not necessarily all of the time period reported in each study. For a few studies that did not clearly define the pandemic period, we defined that period using any indication/reference in the same article for a lockdown or a surge in the number of COVID-19 cases.

Study design label explanations: ‘Same period single year' - Preinterruption measurement at a comparable time period in 2019 only with basic pre-post analysis (unadjusted or adjusted comparison of mean utilisation across the two comparator periods). An example is a study comparing utilisation in the month of March 2020 with utilisation in the month of March 2019; 'Same period multiple years' - Preinterruption measurement at comparable time periods in prior years (2 or more) with basic pre-post analysis.  An example is a study comparing utilisation for weeks 10-16 of 2020 with utilisation during weeks 10-16 in 2019 and 2018 (using the average utilisation from the comparator years) ; 'Time trend single year' – This category refers to studies considering data from an entire year preinterruption time period rather than a single month or period of weeks. An example is a study documenting utilisation for the period January 2019 to some time point in 2020.  In these studies preinterruption utilisation trends may be modelled using data from the prior year to estimate predicted utilisation.  This category also includes studies that do not model prior data but average utilisation across the prior year for comparison to a postinterruption period. An example is a study comparing the monthly average utilisation for the period Jan 1 2019 to Feb 29 2020 with the monthly average utilisation for March in 2020. Both these types of studies would be rated as moderate risk of bias; 'Time trend multiple years' – This category refers to studies considering data from more than one entire year prior to the pandemic interruption. An example is a study documenting utilisation from the period January 2014 to some point in 2020. In these studies preinterruption utilisation trends may be modelled using observations from previous years to estimate utilisation that would have occurred in the absence of the pandemic.

5.2 Table Percentage change in healthcare utilisation for each individual study grouped by category of healthcare utilisation.

| Study | Outcome | Comparator time period* | Weeks being compared | Total volume of services | % Change (95% CI) |
|---|---|---|---|---|---|
| **Admissions** | | | | | |
| **Andersson** | Worsening HF | 2019 | 12 to 13 | 568<br>(C: 353; P: 215) | -30 |
| **Angoulvant** | Ped ED Hospitalisation | 2017/18/19** | 12 to 16 | NR | -45<br>(-32.4 to -57.0) |
| **Athiel** | Gynaecological ED Hospitalisation | 2019 | 10 to 22 | 1761<br>(C: 976; P: 785) | -20 |
| **Baum** | Admissions for any cause | 2019 | 11 to 16 | 130353<br>(C: 85326; P: 45027) | -43<br>(-36.0 to -49.0) |
| **Bollmann** | HF | 2019 | 10 to 18 | 6424<br>(C: 3604; P: 2820) | -21.8<br>(-18.0 to - 26.0) |
| **Bollmann** | Bradycardia | 2019 | 10 to 18 | 624<br>(C: 334; P: 290) | -13.2<br>(-26.0 to +1.0) |
| **Bollmann** | Atrial Fibrillation | 2019 | 10 to 18 | 2962<br>(C: 1640; P: 1322) | -19.4<br>(-13.0 to -25.0) |
| **Bollmann** | Supraventricular tachycardia | 2019 | 10 to 18 | 525<br>(C: 283; P: 242) | -14.5<br>(-28.0 to +1.0) |
| **Bollmann** | Ventricular tachyarrhythmia | 2019 | 10 to 18 | 433<br>(C: 251; P: 182) | -27.5<br>(-13.0 to -40.0) |
| **Braiteh** | ACS | 2019 | 10 to 18 | 180<br>(C: 113; P: 67) | -40.71 |
| **Braiteh** | Admissions for any cause | 2019 | 10 to 18 | 6108<br>(C: 3496; P: 2612) | -25.29 |
| **Claeys** | STEMI | 2017/18/19 | 12 to 14 | NR | -26 |

| | | | | | |
|---|---|---|---|---|---|
| **Clerici** | Psychiatric hospitalisation | 2019 | 8 to 13 | 618 (C: 354; P: 264) | -25.42 |
| **CVD-Covid-UK Consortium** | Total | 2018/19 | 6 to 19 | 1113075 (C: 599372; P: 513703) | -58.2 (-57.5 to -58.9) |
| **De Filippo** | ACS | 2019 | 9 to 13 | 1320 (C: 775; P: 545) | -29.6 (-22.0 to -37.0) |
| **de Havenon** | Stroke | 2018/19 | 6 to 13 | 33867 (C: 17380; P: 16487) | -5.14 |
| **de Havenon** | ACS | 2018/19 | 6 to 13 | 24441 (C: 12111; P: 12330) | 1.81 |
| **De Rosa** | AMI | 2019 | 12 to 19 | 937 (C: 618; P: 319) | -48.4 (-44.6 to-52.5) |
| **De Rosa** | HF | 2019 | 12 to 19 | 236 (C: 154; P: 82) | -46.8 (-39.5 to -55.3) |
| **De Rosa** | Atrial Fibrillation | 2019 | 12 to 19 | 129 (C: 88; P: 41) | -53.4 (-43.9 to -64.9) |
| **De Rosa** | Pulmonary Embolism | 2019 | 12 to 19 | 29 (C: 17; P: 12) | -29.4 (-0.14 to -0.61) |
| **Diegoli** | Stroke | 2019 | 8 to 16 | 1169 (C: 713; P: 456) | -36.15 (-7.7 to -64.6) |
| **Egol** | Hip fracture | 2019 | 6 to 16 | 253 (C: 115; P: 138) | 20 |
| **Enache** | Cardiovascular disease | 2019 | 10 to 13 | 765 (C: 419; P: 346) | -17.42 |
| **Franco** | STEMI | 2019 | 9 to 13 | 215 (C: 105; P: 110) | 4.8 |
| **Franco** | NSTEMI | 2019 | 9 to 13 | 1249 (C: 1105; P: 144) | -87 |
| **Frankfurter** | Worsening HF | 2019 | 10 to 16 | 256 (C: 149; P: 107) | -39.3 (-8.6 to -78.5) |
| **Hoyer** | Strokes admissions | 2019 | 10 to 15 | NR | -15.2 |
| **Hoyer** | TIA admissions | 2019 | 10 to 15 | NR | -38.5 |
| **Jasne** | Strokes admissions | 2019 | 8 to 17 | 863 | -37.2 |

| | | | | | |
|---|---|---|---|---|---|
| | | | | (C: 530; P: 333) | |
| **Kessler** | ACS | 2019 | 10 to 18 | 5920<br>(C: 3411; P: 2509) | -27<br>(-23.0 to -30.0) |
| **Lantelme** | AMI | 2019 | 11 to 14 | 240<br>(C: 142; P: 98) | -30.99 |
| **Li** | STEMI | 2019 | 6 to 18 | 2130<br>(C: 1092; P: 1038) | -4.95 |
| **Mafham** | ACS | 2019 | 2 to 22 | 120076<br>(C: 65375; P: 54701) | -40<br>(-37 to -43) |
| **Manzoni** | Ped | 2019 | 10 to 18 | 91<br>(C: 73; P: 18) | -75 |
| **Onteddu** | Strokes | 2019 | 4 to 20 | 104615<br>(C: 66674; P: 37941) | -43.09 |
| **Papafaklis** | ACS | 2019 | 10 to 15 | 1848<br>(C: 1077; P: 771) | -28.41<br>(-21.0 to -35.0) |
| **Reeves** | STEMI | 2016/17/18/19 | 13 to 17 | 155<br>(C: 85; P: 70) | -17.3 |
| **Reeves** | Stroke | 2016/17/18/19 | 13 to 17 | 230<br>(C: 175; P: 155) | -15.6 |
| **Romaguera** | STEMI | 2019 | 10 to 16 | 919<br>(C: 524; P: 395) | -24.6<br>(-14.0 to -34.0) |
| **Secco** | ACS | 2019 | 10 to 13 | 246<br>(C: 162; P: 84) | -48.15<br>(-33.0 to -61.0) |
| **Seiffert** | Cardiovascular or cerebrovascular emergencies | 2019 | 10 to 22 | 67443<br>(C: 35841; P: 31602) | -14.97 |
| **Zhao** | Stroke | 2019 | 6 to 9 | 56306<br>(C: 34725; P: 21581) | -37.9 |
| **Diagnostics** | | | | | |
| **Collado-Mesa** | Breast imaging | 2018/19 | 14 to 18 | 8239<br>(C: 7142; P: 1097) | -84.64 |
| **Houshyar** | ED volume of all imaging | 2019 | 13 to 14 | 5871<br>(C: 3552; P: 2319) | -34.7<br>(-12.0 to -57.4) |

| | | | | | |
|---|---|---|---|---|---|
| | (MRI, CT, x-ray, US, fluoroscopy) | | | | |
| **Lui** | Upper endoscopies | 2017/18/19 | 4 to 13 | 2700 (C: 1813; P: 887) | -51.1 |
| **Lui** | Lower endoscopies | 2017/18/19 | 4 to 13 | 1681 (C: 1190; P: 491) | -58.7 |
| **Naidich** | Total imaging volume | 2019 | 10 to 16 | 408067 (C: 237388; P: 170679) | -28.1 |
| **Naidich** | ED imaging volume | 2019 | 10 to 16 | 195160 (C: 112579; P: 82581) | -26.6 |
| **Naidich** | Inpatient imaging volume | 2019 | 10 to 16 | 147070 (C: 78902; P: 68168) | -13.6 |
| **Naidich** | Outpatient imaging volume | 2019 | 10 to 16 | 65837 (C: 45907; P: 19930) | -56.6 |
| **Norbash** | All radiological requests | 2019 | 2 to 21 | 282749 (C: 203132; P: 79617) | -21.8 |
| **Toyoda** | Abdominal US | 2018/19 | 6 to 18 | 4506 (C: 2566; P: 1940) | -24.4 |
| **Toyoda** | Abdominal CT/MRIs | 2018/19 | 6 to 18 | 3553 (C: 1874; P: 1679) | -10.38 |
| **Xu** | Optical coherence tomography, indocyanine green, fluorescent angiography | 2018/19 | 11 to 20 | 566955 (C: 355458; P: 211497) | -40.5 (-26.4 to -54.7) |
| **Therapeutics, Procedures, Surgeries** | | | | | |
| **Benazzo** | Trauma surgeries | 2019 | 9 to 14 | 1011 (C: 559; P: 452) | -19.2 |
| **Benazzo** | Femoral neck fracture surgeries | 2019 | 9 to 14 | 656 (C: 349; P: 307) | -12.2 |
| **Bollmann** | Catheter ablations | 2019 | 10 to 18 | 472 (C: 264; P: 208) | -21.2 (-6.0 to -44.0) |
| **Bollmann** | CRM device implantations | 2019 | 10 to 18 | 675 (C: 365; P: 310) | -15.1 (-1.0 to -27.0) |

| | | | | | |
|---|---|---|---|---|---|
| **Bozovich** | Coronary angioplasties | 2019 | 14 to 18 | 1330 (C: 946; P: 384) | -59.41 (-50.0 to -67.0) |
| **Bozovich** | Heart surgeries | 2019 | 14 to 18 | 400 (C: 282; P: 118) | -58.16 (-46.0 to -100) |
| **Bozovich** | PCI | 2019 | 14 to 18 | 2501 (C: 1850; P: 651) | -64.81 (-50.0 to -78.0) |
| **Bozovich** | General surgeries | 2019 | 14 to 18 | 24805 (C: 19600; P: 5205) | -73.44 (-62.0 to -75.0) |
| **Bozovich** | Chemotherapy and radiotherapy | 2019 | 14 to 18 | 9227 (C: 5005; P: 4222) | -15.64 (-3.0 to -52.0) |
| **Bozovich** | GI endoscopies | 2019 | 14 to 18 | 8549 (C: 7137; P: 1412) | -80.22 (-77.0 to -93.0) |
| **Bramer** | Non-influenza immunisation for children | 2017/18/19 | 1 to 18 | NR | -21.5 |
| **Cano-Valderrama** | Acute surgeries | 2019 | 12 to 17 | 402 (C: 285; P: 117) | -58.95 |
| **de Havenon** | MT | 2018/19 | 6 to 13 | 725 (C: 319; P: 406) | 27.3 |
| **de Havenon** | tPA | 2018/19 | 6 to 13 | 570 (C: 266; P: 304) | 14.3 |
| **de Havenon** | PCI | 2018/19 | 6 to 13 | 2596 (C: 1330; P: 1266) | -4.81 |
| **Garcia** | Cardiac catheterisation | 2019 | 10 to 18 | 1332 (C: 779; P: 553) | -29.1 |
| **Gawron** | Gastrointestinal endoscopies | 2019 | 10 to 18 | 34053 (C: 23455; P: 10598) | -54.81 |
| **Gawron** | Colonoscopies | 2019 | 10 to 18 | 57183 (C: 43371; P: 13812) | -68.15 |
| **Giuntoli** | Scheduled orthopaedic procedures | 2019 | 10 to 13 | 583 (C: 444; P: 139) | -68.69 |

| | | | | | |
|---|---|---|---|---|---|
| **Giuntoli** | Trauma orthopaedic procedures | 2019 | 10 to 13 | 488<br>(C: 270; P: 218) | -19.26 |
| **Gruttadauria** | Liver transplantation and related procedures | 2018/19 | 10 to 11 | 98<br>(C: 61; P: 37) | -39.34 |
| **Kadavath** | Invasive cardiac procedures | 2019 | 10 to 16 | 7219<br>(C: 4671; P: 2548) | -45.45 |
| **Kerleroux** | MT for stroke | 2019 | 8 to 13 | 1512<br>(C: 844; P: 668) | -21<br>(-18.0 to -24.0) |
| **Langdon-Embry** | Routine childhood immunisation | 2019 | 12 to 22 | 590000<br>(C: 344000; P: 246000) | -28.49 |
| **Lazaros** | Cardiac surgery procedures | 2019 | 12 to 19 | 330<br>(C: 246; P: 84) | -65.85 |
| **Mafham** | PCI after the admission day | 2019 | 2 to 22 | 17469<br>(C: 8055; P: 9414) | -47<br>(-37 to -52) |
| **Mafham** | PCI on the admission day | 0 | 2 to 22 | 19277<br>(C: NR; P: NR) | -16<br>(-7 to -24) |
| **Mafham** | CABG | 2019 | 2 to 22 | 3196<br>(C: 2663; P: 533) | -80<br>(-68 to -87) |
| **Mafham** | Angiography | 2019 | 2 to 22 | 16079<br>(C: 11485; P: 4594) | -60<br>(-53 to -65) |
| **Mazzatenta** | Non-urgent surgical procedures | 2018/19 | 12 to 15 | 918<br>(C: 713; P: 205) | -71.25 |
| **Mazzatenta** | Urgent surgical procedures | 2018/19 | 12 to 15 | 274<br>(C: 161; P: 113) | -29.6 |
| **McDonald** | Hexavalent vaccine (first does) | 2019 | 10 to 17 | 62692<br>(C: 31475; P: 31217) | -0.82 |
| **McDonald** | MMR vaccine (first does) | 2019 | 10 to 17 | 59809<br>(C: 30989; P: 28820) | -7 |
| **Onteddu** | tPA | 2019 | 4 to 20 | 1841 | -50.93 |

| | | | | | |
|---|---|---|---|---|---|
| | | | | (C: 1235; P: 606) | |
| **Onteddu** | MV | 2019 | 4 to 20 | 644 (C: 399; P: 245) | -38.6 |
| **Pinar** | Urological surgeries | 2019 | 12 to 13 | 1439 (C: 995; P: 444) | -55.4 |
| **Polo Lopez** | Congenital heart diseases surgeries | 2019 | 12 to 20 | 193 (C: 142; P: 51) | -51 |
| **Range** | Coronary angiography for STEMI | 2019 | 10 to 13 | 430 (C: 246; P: 184) | -25.2 |
| **Requena** | Frozen embryo transfer | 2019 | 6 to 12 | 4461 (C: 2500; P: 1961) | -21.5 |
| **Requena** | IVF | 2019 | 6 to 12 | 5441 (C: 3007; P: 2434) | -19.1 |
| **Requena** | IUI | 2019 | 6 to 12 | 1301 (C: 564; P: 467) | -17.3 |
| **Salerno** | Urgent GI endoscopic procedures | 2019 | 10 to 13 | 2305 (C: NR; P: NR) | -39.49 |
| **Tinay** | Non-deferrable uro-oncological procedures | 2019 | 11 to 15 | 290 (C: 200; P: 90) | -55 |
| **Wong** | Orthopaedic operations | 2016/17/18/19 | 5 to 13 | 928278 (C: 595814; P: 332464) | -44.2 (-54.7 to -33.7) |
| **Xu** | Intravitreal injections | 2018/19 | 11 to 20 | 454765 (C: 235996; P: 218769) | -7.3 (2.2 to -16.8) |
| **Zhao** | Thrombolysis | 2019 | 6 to 9 | 5930 (C: 3422; P: 2508) | -25.5 |
| **Zhao** | Thrombectomy | 2019 | 6 to 9 | 2268 (C: 1298; P: 970) | -22.7 |
| **Visits** | | | | | |
| **Abdulmalik** | All primary care services | 2018/19 | 10 to 22 | 1384037 (C: 872691; P: 511346) | -41.41 |

| | | | | | |
|---|---|---|---|---|---|
| **Angoulvant** | Ped ED | 2017/18/19** | 12 to 16 | 871543<br>(C: NR; P: NR) | -68<br>(-55.8 to -81.2) |
| **Antonucci** | ED urological | 2019 | 10 to 18 | 304<br>(C: 201; P: 103) | 48.8 |
| **Athiel** | Gynaecological ED | 2019 | 10 to 22 | 39690<br>(C: 24982; P: 14708) | -41 |
| **Bayles, preprint** | ED | 2018/19 | 12 to 18 | 21527<br>(C: 17230; P: 4297) | -50.1<br>(-39.5 to -60.7) |
| **Benazzo** | Orthopaedic outpatient | 2019 | 9 to 14 | 17041<br>(C: 6863; P: 10178) | -48.3 |
| **Benazzo** | ED trauma | 2019 | 9 to 14 | 14772<br>(C: 6050; P: 8722) | -44.17 |
| **Benazzo** | Elective orthopaedic surgeries | 2019 | 9 to 14 | 8113<br>(C: 3065; P: 5048) | -64.7 |
| **Bozovich** | ED | 2019 | 14 to 18 | 268899<br>(C: 213947; P: 54952) | -74.32<br>(-65.0 to -79.0) |
| **Butt** | ED | 2019 | 10 to 18 | 102033<br>(C: 58858; P: 43175) | -26.7 |
| **Cheek** | ED | 2019 | 13 to 21 | 41041<br>(C: 26871; P: 14170) | -47.27<br>(-44.2 to -50.3) |
| **Chou** | Hospice home care visits | 2019 | 1 to 18 | 1516<br>(C: 777; P: 739) | -4.89 |
| **CVD-Covid-UK Consortium** | ED | 2018/19 | 6 to 19 | 942169<br>(C: 506516; P: 435653) | -52.8<br>(-52.2 to -53.5) |
| **CVD-Covid-UK Consortium** | ED cardiac | 2018/19 | 6 to 19 | NR | -40.2<br>(-35.6 to -45.0) |
| **CVD-Covid-UK Consortium** | ED cerebrovascular | 2018/19 | 6 to 19 | NR | -31.8<br>(-26.2 to -38.0) |

| | | | | | |
|---|---|---|---|---|---|
| **CVD-Covid-UK Consortium** | ED vascular | 2018/19 | 6 to 19 | NR | -40.6 (-31.5 to -50.3) |
| **Frankfurter** | Symptoms suggestive of HF | 2019 | 10 to 16 | 1906 (C: 800; P: 1106) | 38.3 (26.3 to 51.6) |
| **Frankfurter** | HF | 2019 | 10 to 16 | 314 (C: 186; P: 128) | -43.5 (-14.8 to -79.4) |
| **Giuntoli** | Orthopaedic first aid visits | 2019 | 10 to 13 | 1679 (C: 1301; P: 378) | -70.95 |
| **Hartnett** | ED | 2019 | 11 to 22 | 3319945 (C: 2099734; P: 1220211) | -31.47 |
| **Isba** | Ped ED | 2019 | 6 to 13 | NA (C: NA; P: NA) | -17.74 |
| **Kim** | ED | 2019 | 11 to 18 | 68384 (C: 38712; P: 29672) | -44 (-33.0 to -53.0) |
| **Kolbaek** | Referrals to psychiatric services | 2019 | 9 to 18 | 7982 (C: 4419; P: 3563) | -19.4 |
| **Krenzlin** | ED Neurosurgery | 2018/19 | 12 to 16 | 2646 (C: 1824; P: 822) | -44.7 (-42.6 to -46.8) |
| **Lazzerini** | Ped ED | 2019 | 10 to 13 | 10826 (C: 8818; P: 2008) | -77.72 (-73.0 to -88.0) |
| **Manzoni** | Ped ED | 2019 | 10 to 18 | 1654 (C: 1428; P: 226) | -86 (-32.0 to -55.0) |
| **Mazzatenta** | Outpatient neuro-surgical | 2018/19 | 12 to 15 | 2234 (C: 1768; P: 466) | -73.6 |
| **Mitchell** | ED | 2017/18/19 | 14 to 17 | 14059 (C: 8643; P: 5416) | -37.3 (-33.0 to -41.0) |
| **Novara** | ED urological | 2019 | 12 | 399 (C: 275; P: 124) | -54.9 |
| **Pignon** | ED psychiatric | 2019 | 12 to 15 | 1777 (C: 1224; P: 553) | -54.8 |
| **Pop** | Stroke | 2019 | 10 to 13 | 462 | -39.6 |

| | | | | | |
|---|---|---|---|---|---|
| | | | | (C: 288; P: 174) | |
| **Qasim** | Trauma | 2019 | 11 to 16 | 2386<br>(C: 1328; P: 1058) | -20.3 |
| **Santana** | ED | 2019** | 10 to 13 | 863414<br>(C: NR; P: NR) | -47.98 |
| **Scaramuzza** | Ped ED | 2019 | 9 to 13 | 3912<br>(C: 2958; P: 954) | -67.8 |
| **Scholz** | STEMI | 2017/18/19 | 10 to 13 | 1716<br>(C: 1329; P: 387) | -12.64 |
| **Smalley** | ED | 2019 | 13 to 17 | 87840<br>(C: 56453; P: 31387) | -44.4 |
| **Toro** | ED | 2015/16/17/18/19 | 10 to 18 | 5045647<br>(C: 3198508; P: 1847139) | -42.25 |
| **Toro** | Circulatory system ED | 2015/16/17/18/19 | 10 to 18 | 105471<br>(C: 58439; P: 47032) | -19.52 |
| **Toro** | Stroke ED | 2015/16/17/18/19 | 10 to 18 | 11004<br>(C: 6385; P: 4619) | -27.66 |
| **Toyoda** | Liver clinics | 2018/19 | 6 to 18 | 8568<br>(C: 5335; P: 3233) | -39.4 |
| **Xu** | Retinal outpatient clinics | 2018/19 | 11 to 20 | 813585<br>(C: 485433; P: 328152) | -32.4<br>(-20.4 to - 44.4) |

*this is the comparator year that studies included in their comparison to the 2020 time period; **these studies compared the expected/forecasted utilisation for 2020 from data from these years

Abbreviations: ED: emergency department; HF: Heart Failure; IVF: In vitro fertilisation; IUI: Intrauterine insemination; MT: Mechanical thrombectomy; tPA: tissue Plasminogen Activator; CABG: Coronary artery bypass grafting; ACS: Acute Coronary Syndrome; AMI: Acute Myocardial Infarction; STEMI: ST Elevation Myocardial Infarction; MRI: Magnetic Resonance Imagine; CT: computerized tomography; US: Ultrasonography; CRM: Cardiac rhythm management; PCI: Percutaneous Coronary Interventions; GI: Gastrointestinal

For studies that reported the changes in healthcare services as incidence rate ratios, IRR, we estimated the % change in healthcare services as 100* (1-IRR). For example, IRR of 0.75 converted to 25% reduction in healthcare services

5.3 Table of results of secondary outcomes of the included studies

| Study | Secondary Outcome | Change in proportions of severe patients* | P-value, if provided |
|---|---|---|---|
| **Andersson** | Mortality | No change | 0.45 |
| **Braiteh** | STEMI/NSTEMI | No change | NR |
| **Butt** | % ACS from those presented with cardiac symptoms | Increase | NR |
| **Cano-Valderrama** | SOFA score >0 | No change | 0.16 |
| **Claeys** | % Cardiac arrest | No change | 0.7 |
| **Claeys** | Killip class | No change | 0.7 |
| **Claeys** | Mortality | No change | 0.6 |
| **Clerici** | Voluntary/involuntary admission | Increase | NR |
| **Collado-Mesa** | Positive biopsy (diagnostic yield) | No change | NR |
| **CVD-COVID** | Procedures for cardiac, cerebrovascular, other vascular conditions | No change | NR |
| **De Rosa** | Mortality | Increase | <0.001 |
| **De Rosa** | STEMI/NSTEMI | Increase | NR |
| **De-Filippo** | STEMI/NSTEMI | No change | 0 |
| **Diegoli** | Admissions for severe stroke (NIH stroke scale score) | Increase | NR |
| **Egol** | Mortality (In-patient and 30 day) | Increase | 0.005-0.035 |
| **Egol** | Non-operative cases | No change | 0.793 |
| **Frankfurter** | Hospitalisation | No change | 0.22 |
| **Frankfurter** | ICU admission | No change | 0.86 |
| **Frankfurter** | In-hospital mortality | No change | 0.05 |
| **Frankfurter** | NYHA class III-IV | No change | 0.3 |
| **Giuntoli** | Hospitalisation | Increase | NR |
| **Hoyer** | Stroke/TIA | Increase | NR |
| **Kerleroux** | % unwitnessed onset | Increase | 0.004 |
| **Kerleroux** | ASPECTs score | Increase | 0.041 |

| | | | |
|---|---|---|---|
| **Kerleroux** | Baseline NIHSS | No change | 0.279 |
| **Kessler** | STEMI/NSTEMI | No change | 0 |
| **Lazaros** | Emergency/nonemergency | Increase | <0.001 |
| **Lui** | Positive rate for colon cancer | Increase | <0.001 |
| **Lui** | Positive rate for gastric cancer | No change | 0.14 |
| **Mafham** | STEMI/NSTEMI | Increase | NR |
| **Manzoni** | Hospitalisation | Increase | <0.001 |
| **Mazzatenta** | Urgent/Nonurgent | Increase | NR |
| **Mitchell** | Triage category | No change | NR |
| **Novara** | Hospitalisation | No change | 0.8 |
| **Novara** | Triage category | No change | 0.06 |
| **Papafakis** | STEMI/NSTEMI | Increase | NR |
| **Pignon** | Hospitalisation | No change | 0.872 |
| **Pop** | admission | Increase | NR |
| **Pop** | Initial NIHSS score | No change | 0.886 |
| **Qasim** | Changes in % of all trauma volume that was at the highest level of acuity (described as 'alert') | Increase | 0.006 |
| **Romaguera** | % of patients with sudden cardiac death | No change | 0 |
| **Romaguera** | 10-day mortality | No change | 0.459 |
| **Romaguera** | Killip class II-IV | No change | 0.8 |
| **Salerno** | Diagnostic yield for urgent EGDs | Increase | <0.001 |
| **Salerno** | Diagnostic yield for urgent lower endoscopy | No change | 0.3 |
| **Santana** | Triage category | No change | 0 |
| **Scaramuzza** | Triage category | Increase | 0 |
| **Scholz** | In-hospital mortality | No change | 0.68 |
| **Scholz** | TIMI score | No change | 0.464 |
| **Secco** | GRACE score | Increase | <0.01 |
| **Secco** | Peak troponin | Increase | <0.01 |
| **Secco** | STEMI/NSTEMI | Increase | <0.01 |
| **Secco** | Mortality | No change | NS |

| | | | |
|---|---|---|---|
| **Seiffert** | Acute stroke/TIA | Increase | 0 |
| **Seiffert** | STEMI/NSTEMI | Increase | 0 |
| **Seiffert** | In-hospital mortality | No change | 0 |
| **Seiffert** | Intervention/surgeries | No change | 0 |
| **Tinay** | ASA scores | Increase | 0.005 |
| **Toyoda** | Visits in advanced disease patients | No change | 0.11 |
| **Wong** | Emergency/elective | Increase | NR |

Note: *This secondary outcome domain is exploring, if there is a reduction in services, whether or not there is a greater or lesser reduction in the proportion of patients/people using the service who have milder or more severe forms of illness. If there is an increase in the proportions with more severe illness - which means a greater reduction among those with milder illness – then an "increase" is recorded in this column.

#### 5.4 Figures Change in healthcare utilisation for each category of healthcare services:

Each dot represents a study estimate for each calendar week. For studies that only provided averages of changes for the whole study period, we plotted the average estimates for each calendar week of the corresponding study period.

Figure 5.4a visits

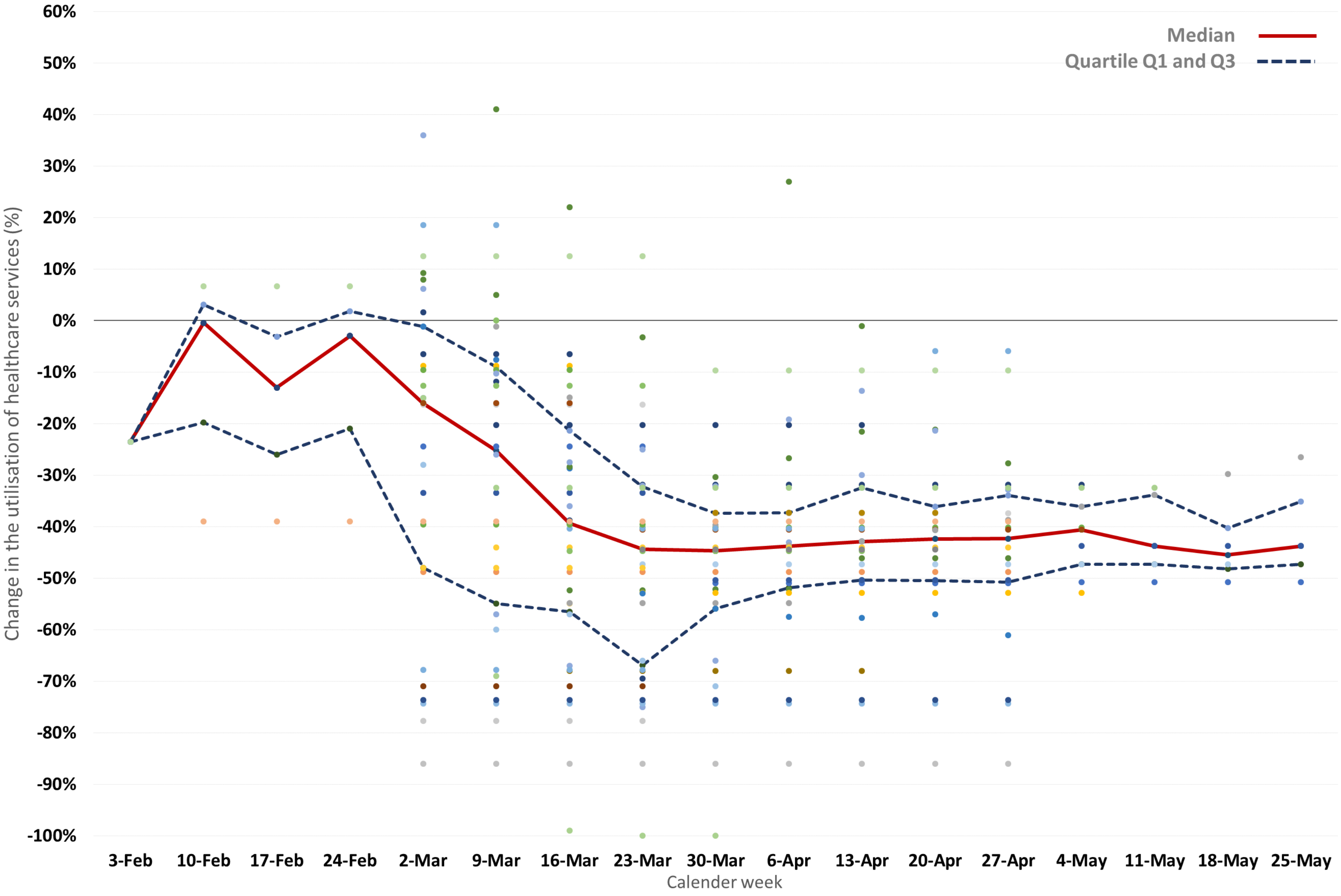

**Figure 5.4b admissions**

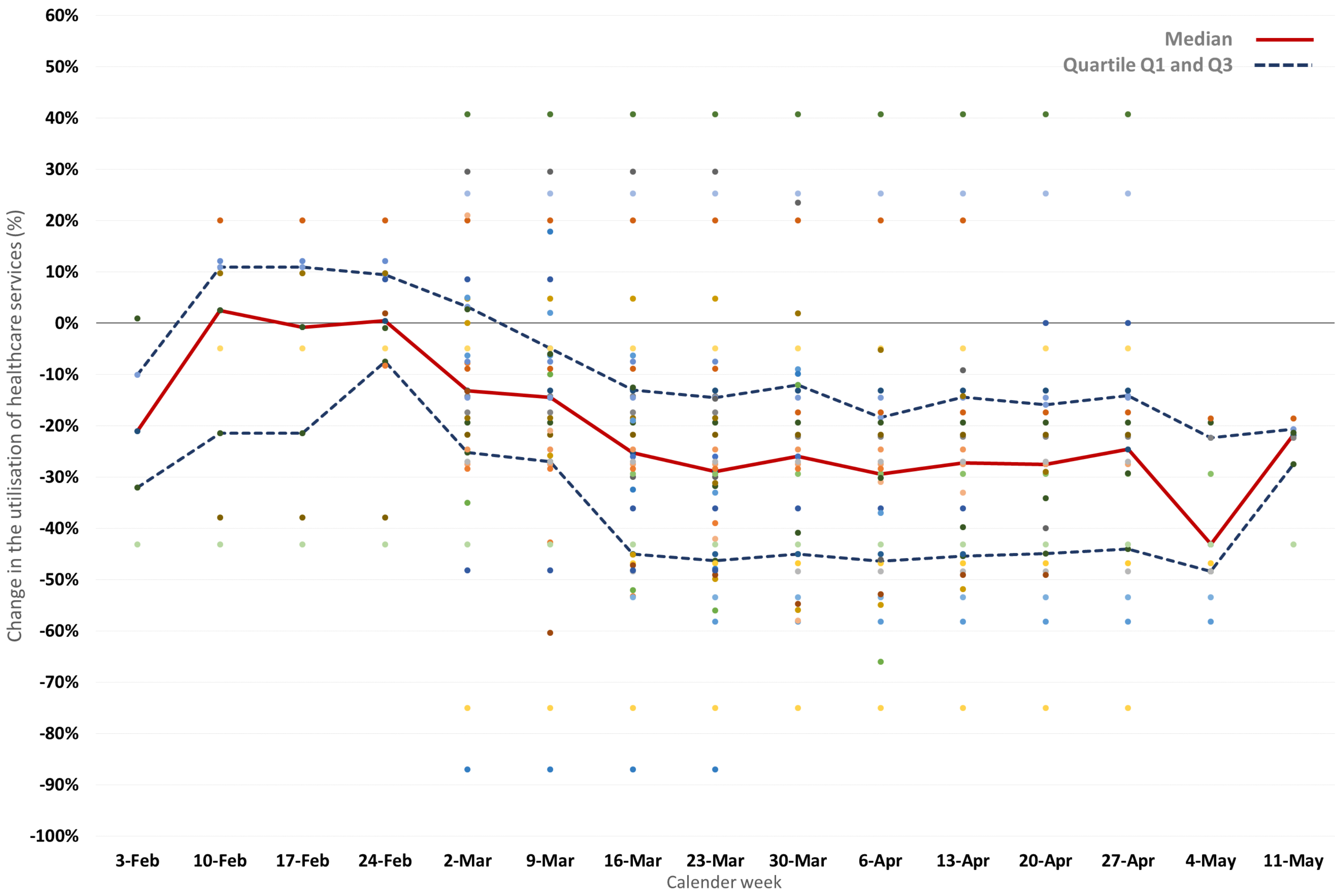

**Figure 5.4c diagnostics**

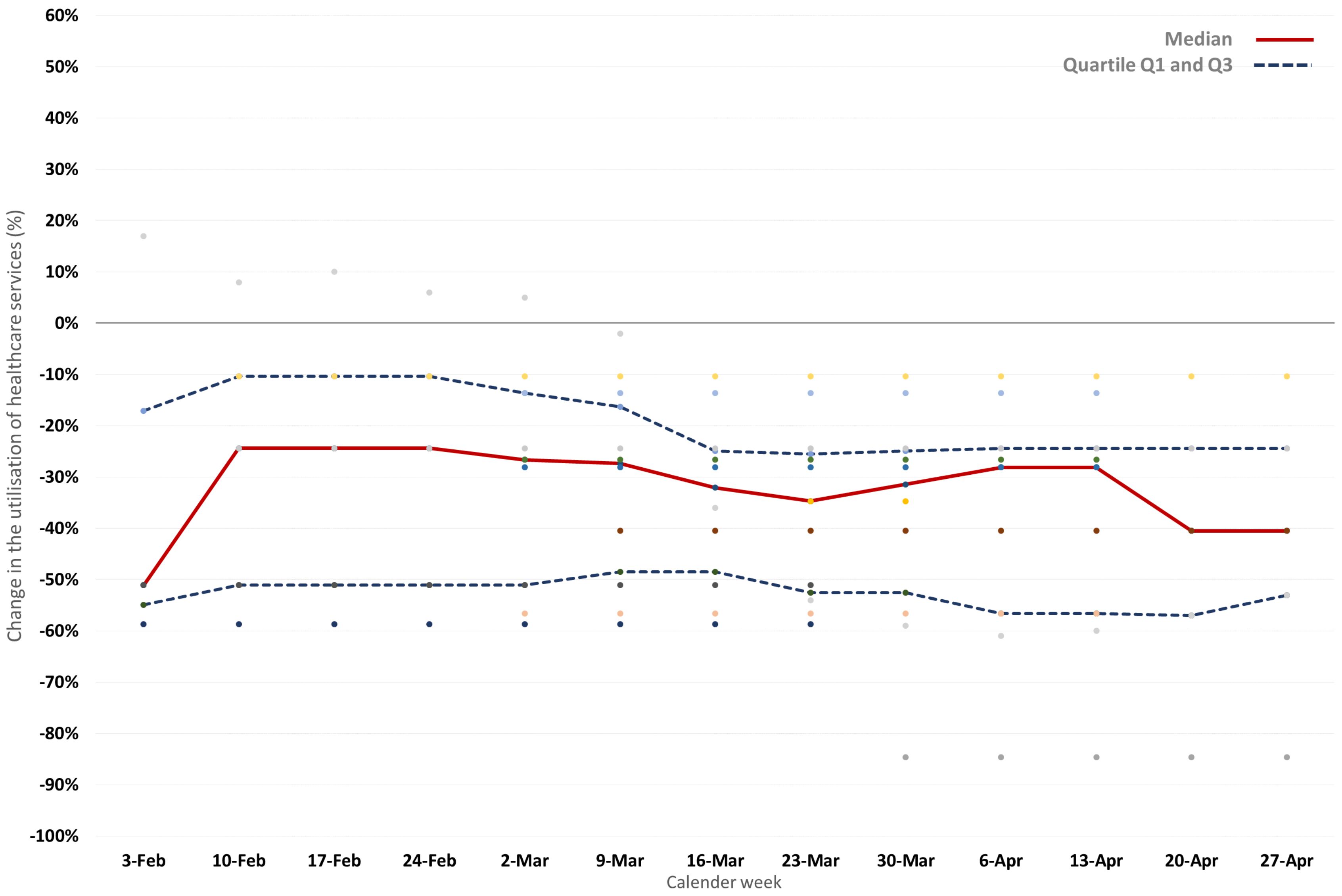

**Figure 5.4d therapeutics**

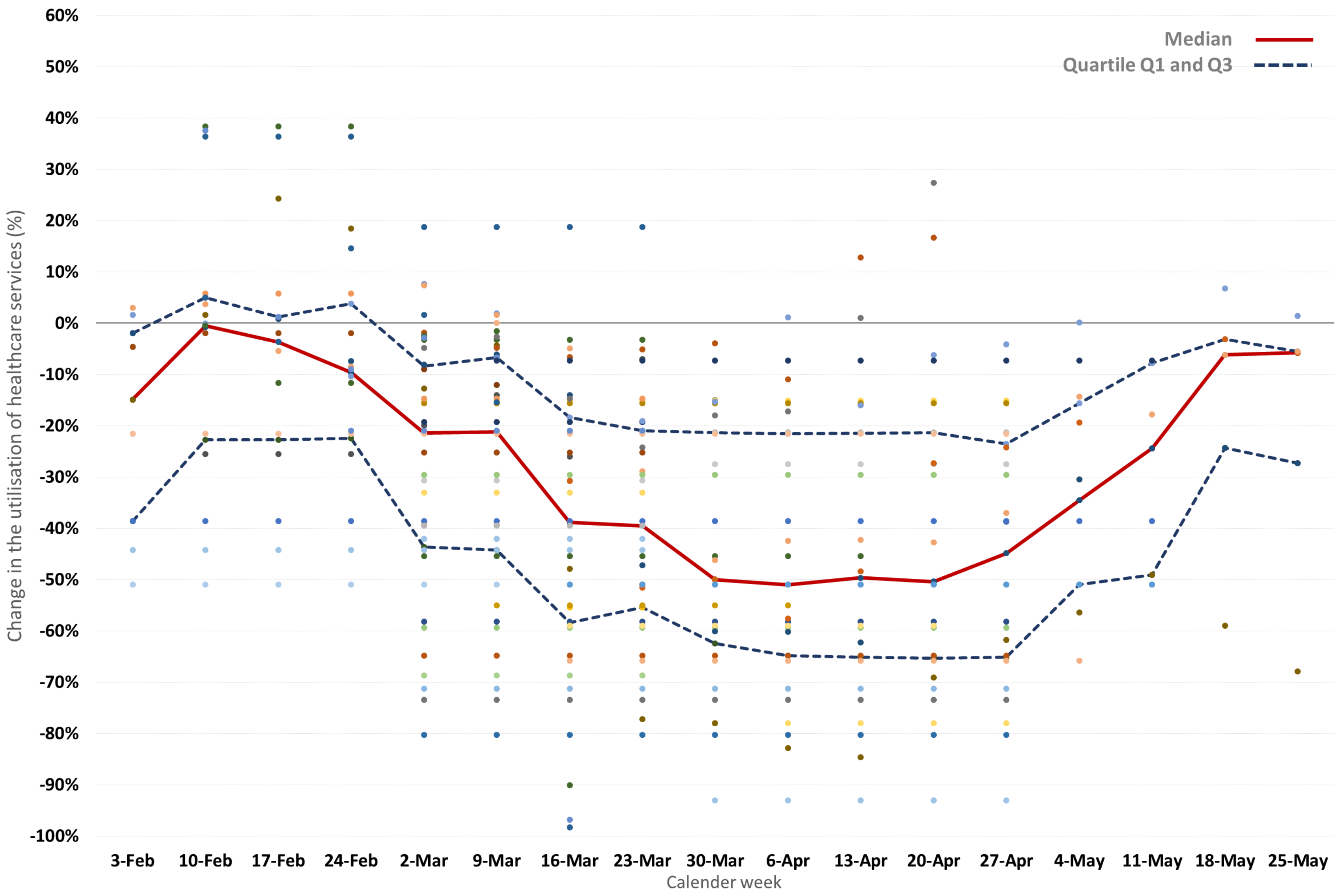
